# Don’t Jump to Faulty Conclusions: Using the Synthetic Control Method to Evaluate the Effect of a Counterfactual Lockdown in Sweden

**DOI:** 10.1101/2024.09.20.24314059

**Authors:** Jonas Herby

## Abstract

Several studies based on the Synthetic Control Method (SCM) link the many COVID-19 deaths in Sweden during the spring of 2020 to its decision not to lock down. These studies predominantly rely on a limited pre-intervention period and tend to focus on the very short run neglecting that Swedish health authorities emphasized that their public health strategy was designed for the long run. This paper handles these shortcomings expanding both the pre- and post-intervention period considerably using weekly mortality rates. I find no significant effect of the absence of lockdown on COVID-19 mortality neither in the short or long run. Using the timing of the winter holiday as a proxy for the extent/spread of COVID-19 in societies before lockdown – a variable which is unobservable due to limited testing – I suggest that the effect of lockdowns found in earlier studies using Sweden as a control are likely to be partially driven by unobserved variables rather than policies.

## Introduction

In the spring of 2020, Sweden, led by state epidemiologist Anders Tegnell, opted against a mandatory lockdown, a decision that drew significant international attention and academic scrutiny. Despite reporting low excess mortality overall during the pandemic, Sweden experienced substantial COVID-19 related deaths early on.

The case of Sweden has fueled a body of research, and several studies have tried to estimate the effect of lockdowns using Sweden as a control group. These studies, which mostly focus on the first pandemic wave, generally conclude that a Swedish lockdown would have prevented thousands of deaths in Sweden in the spring of 2020.

In general, there are two groups of studies examining the effect of lockdowns using Sweden as a control group. The first group compares Sweden to a relatively small control group using a difference-in-difference approach. For example, Gordon et al. (2020) compare Sweden to its Nordic neighbors and New Zealand (SIC!) and conclude that “imposition of full international travel restrictions combined with high levels of government mandated stringency of SD [social distancing] reduced the per capita cases and per capita fatalities associated with COVID-19 in 2020”. Conyon et al. (2020) compare Sweden to Norway and Denmark and conclude that “stricter lockdown policies are associated with fewer COVID-19 deaths”, and Juranek and Zoutman (2021) compare Denmark and Norway to Sweden and conclude that “in the absence of strict NPIs […], Denmark (Norway) would have had 402 (1015) percent more deaths”. The limited number of observations in these studies do not permit the use of advanced econometric techniques to handle potential biases.

The second group of studies employs the Synthetic Control Method (SCM). The SCM is a statistical technique used to estimate the causal effects of interventions in observational studies, particularly when randomized control trials are not feasible. This method constructs a synthetic version of the treated unit by optimally weighing a combination of units from a donor pool that did not receive the intervention but share similar pre-intervention characteristics and outcomes. The goal of SCM is to create a counterfactual scenario that closely approximates what would have happened to the treated unit in the absence of the intervention, thereby isolating the intervention’s effect on the outcome of interest.

Based on a 24 day pre-intervention window, Conyon and Thomsen (2021) conclude that “stricter lock-down policies are associated with fewer COVID-19 infections and deaths”, and Cho (2020) uses a 25 day pre-intervention period and find that “lockdown measures would have been associated with a lower excess mortality rate in Sweden by 25 percentage points”. Born et al. (2021) employ a 16 day pre-intervention period and find that “a 9-week lockdown in the first half of 2020 would have reduced […] deaths by about 38%”, and Latour et al. (2022) find that “a lockdown would have had sizable effects within one week” based on a 15 day pre-intervention window. Recently Ege et al. (2023) found that “not imposing a mandatory lockdown resulted in […] a substantial increase in mortality” using excess mortalities over a 20 week pre-intervention window.

The SCM method has also been applied in other locations, but Herby et al. (2023) show that these studies tend to focus on outliers with notably high COVID-19 mortality rates during the first wave. Herby et al. (2023) report that at least seven out of eleven eligible SCM studies targeted regions that experienced substantial deaths early in the pandemic.^1^ In contrast, Mader and Rüttenauer (2021), who use the Generalized Synthetic Control Method (GSCM) covering 169 countries to avoid selection bias, ‘do not find substantial and consistent mitigating effects of any NPI under investigation on COVID-19-related deaths per capita.’

While the effects of lockdown estimated in the SCM-studies are significantly smaller than those predicted by epidemiological studies, such as those by Ferguson et al. (2020) – with most estimates suggesting the number of avoided deaths to be in the lower thousands for a population with the size of that of Denmark^2^ – they are also constrained by very short pre-intervention windows.

In this study I qualify the use of the SCM in relation to examining the effect of a lockdown in Sweden in three ways.

First, I use mortality rates to extend the pre-intervention period considerably compared to above-mentioned studies. Abadie (2021) underscores the significance of a comprehensive pre-intervention period when designing synthetic control studies, stating that “it is of crucial importance to collect information on the affected unit and the donor pool for a large pre-intervention window”. The term “large pre-intervention window” is inherently ambiguous; however, asserting that a pre-intervention window of merely a few weeks qualifies as “large” is, under any circumstances, a contention that warrants critical examination.

Second, I extend the post-intervention period considerably compared to above-mentioned studies. Through the pandemic, Swedish health authorities emphasized that their public health strategy was designed for the long run. One key reason for avoiding strict lockdowns was to prevent fatigue among the population, which could potentially lead to higher mortality rates over time. This approach can be interpreted as a strategic trade-off, accepting a higher number of deaths in the short run to reduce the risk of greater mortality in the future. Thus, evaluating the Swedish COVID-19 policy on the short run may lead to wrong conclusions.

Third, I examine the impact of the timing of the winter holiday. Arnarson (2021) and Björk et al. (2021) illustrate how the timing of the winter holiday significantly influenced the spread of the virus in Europe. Their research indicates that regions which scheduled their winter holidays later, in weeks 9 or 10 rather than weeks 6 to 8, experienced disproportionately high COVID-19 case numbers during the initial wave.^3^ This was largely due to virus transmission by ski tourists returning from the Alps. Specifically, regions with their winter holiday in week 9 had, ceteris paribus, almost twice (90%) the COVID-19 cases in March 2020 compared to those with winter holiday in week 7.

In Sweden, where approximately one-third of the population, including the heavily affected Stockholm region, had their winter holiday in week 9, this timing presents a critical omitted variable for studies evaluating the effectiveness of non-lockdown approaches. Many Swedes traveling to the Alps during this period were exposed to a substantial yet undetected outbreak. This unique circumstance, combined with Sweden’s decision against a lockdown, complicates direct comparisons with other nations and highlights the necessity for nuanced analyses that account for such significant omitted variables.

### Using SCM to assess effects

I find that a counter factual lockdown would have reduced mortality rates by approximately 18 deaths per 100,000 in the short run (by week 26 of 2020), but *increased* mortality by more than 25 deaths per 100,000 in the longer run (week 26 of 2021). Crucially, both of these results are far from significant.

I also find large discrepancies in the effect estimates across regions with different timing of their winter holiday suggesting that differences in excess mortality are less a result of the no-lockdown policy and more a result of unobserved variables such as the extent/spread of COVID-19 before lockdown. This result indicates that the effect of lockdowns found in other studies using Sweden as a control are at least partially driven by unobserved variables too.

## Data & Methodology

### Methodology

The SCM is a specialized variant of the difference-in-difference analysis which serves as an econometric tool for policy evaluation. Noted for its interpretability and transparency, SCM is particularly valuable in constructing counterfactual scenarios to assess the impact of interventions without randomized control trials, cf. Abadie (2021). In this study, SCM is employed to create a “synthetic” (also called “doppelgänger”), which hypothetically adheres to lockdown measures. This enables a robust comparative analysis between the actual pandemic trajectories observed in Sweden and those that would have potentially occurred under lockdown conditions.

The construction of the synthetic control involves selecting a donor pool of potential control units (countries) and then determining a set of non-negative weights that minimize the difference between the treated unit and the synthetic control in terms of pre-intervention characteristics and outcomes. This weighted combination forms a synthetic control that serves as a benchmark for comparison during the post-intervention period. The causal effect of the intervention is estimated by comparing the post-intervention outcomes of the treated unit with those of the synthetic control.

SCM has been widely applied in various fields, including economics, political science, and public health, due to its ability to handle complex longitudinal data and provide transparent and intuitive results. For instance, Abadie and Gardeazabal (2003) utilized SCM to assess the economic impact of terrorism in the Basque Country by comparing it with a synthetic counterpart composed of other Spanish regions. Similarly, Abadie et al. (2010) employed this method to evaluate the effects of California’s tobacco control program by constructing a synthetic control from other U.S. states. Additionally, Grier and Maynard (2016) used SCM to analyze the economic consequences of Hugo Chavez’s policies in Venezuela, creating a synthetic control from similar countries to isolate the impact of Chavez’s administration on Venezuela’s economy, demonstrating significant negative effects.

Ege et al. (2023) use cumulative excess mortality to construct their synthetic controls. However, this approach raises several issues. First, there is little reason to believe that the initial spread of COVID-19 during the early days of the pandemic correlates with cumulative excess mortality. Even if deaths from influenza or other respiratory viruses in a typical winter season spread to Europe in a way similar to COVID-19 (e.g. via ski tourists in the Alps, as Arnarson (2021); Björk et al. (2021) show was the case with COVID-19), this would manifest itself in data as mortality, not (cumulative) excess mortality. Second, since the COVID-19 pandemic in Europe began in March, it is not comparable to earlier seasons where mortality typically peaks months before COVID-19 deaths peaked. Third, using cumulative excess mortality makes the synthetic control calculation very sensitive to the specific starting point of the analysis, specifically the choice of when the cumulative count is set to zero. Figure 11 in Appendix A shows that the results in Ege et al. (2023) change dramatically, if one set the zero point just one week later.

Hence, I construct my synthetic controls based on mortality rates. Thus, my synthetic control will illustrate expected mortality in the post-intervention window (and not expected excess deaths as in Ege et al. (2023)) and the difference between actual Sweden and the synthetic control can therefore be interpreted as the excess mortality.

Besides mortality rates, I also employ hospital beds per 1,000 people, urban population as share of total population, GDP per capita (PPP), share of population aged 65 and above, and the migrant share of the population as control variables.

### Data

I employ three primary data sources.

First, I source weekly mortality data for donor countries from the Short-Term Mortality Fluctuations (STMF) data provided by mortality.org (accessed July 2, 2024). The STMF collection is derived from official national statistics and adjusted for completeness and consistency, ensuring a high level of reliability. This comprehensive dataset facilitates cross-national comparisons and temporal analyses that are essential for public health planning, policy-making, and epidemiological research. I calculate the weekly mortality rates as the yearly mortality rate in STMF (variable “RTotal”) divided by 52.^4^

Second, I source data for hospital beds per 1,000 people, urban population as share of total population, GDP per capita (PPP), share of population aged 65 and above, and the migrant share of the population from the World Bank’s World Development Indicators database (accessed June 30, 2024).^5^ I use simple forward/backward fill to deal with missing data.

Third, I utilize weekly deaths and population data from SCB Statistics Sweden for detailed regional analysis within Sweden (accessed June 30, 2024).^6^ Importantly, the data from SCB Statistics for the entirety of Sweden corresponds precisely with the mortality data provided by mortality.org, ensuring consistency and accuracy in the analysis.

## Extending the scope of SCM in relation to evaluating Sweden’s COVID-19 policy

I expand on three critical aspects of the SCM, which have significant implications for evaluating the consequences of Sweden’s decision not to implement lockdown measures, as well as for interpreting results from other studies that employ similar methodologies.

### The impact of a short pre-intervention window

Most existing SCM studies examining Sweden’s COVID-19 policy are based on very short pre-intervention – often just a few weeks long. This approach appears to contrast sharply with the guidelines suggested by Abadie (2021), who emphasized the importance of using a “large pre-intervention window” to gather sufficient data on the affected unit and the donor pool. Compared to previous studies, by using mortality rates from mortality.org, I can extend the pre-intervention period considerably, as STMF, SCB and the World Bank supply data for 26 countries going back to (at least) 2007 and 36 countries going back to 2016.^7^

### The impact of a short post-intervention window

The mentioned SCM studies all end their analysis in the fall of 2020 focusing solely on the first part of the pandemic. While some studies are limited by data availability at the date of publication, Ege et al. (2023) ends their analysis arguing that “by November, synthetic Sweden is no longer an appropriate counterfactual for Sweden due to policy changes in the donor countries.” I see two potential flaws in this argument.

First, although Sweden did implement more mandatory measures during the 2020/2021 winter the restrictions were still much more lenient than in other (donor) countries. For example, Sweden didn’t close elementary schools, and although there were restrictions on bars and restaurants they were not closed down. Hairdressers, fitness centers, shopping malls etc. were also open, and curfews were never implemented.

Second, Swedish health authorities emphasized that their public health strategy was designed for the long run. One key reason for avoiding strict lockdowns was to prevent fatigue among the population, which could potentially lead to higher mortality rates over time. This approach can be interpreted as a strategic trade-off, accepting a higher number of deaths in the short run to reduce the risk of greater mortality in the future. Supporting this perspective, Mulligan and Arnott (2022) have demonstrated that non-COVID excess deaths increased following the implementation of lockdowns. Consequently, a short-term focus on pandemic management may not be the most appropriate course of action to understand the effect of lockdowns.

### The impact of the timing of the winter holiday

The timing of the winter holiday emerges as a substantial factor in the spread of COVID-19, particularly among regions that observed their winter holidays later (weeks 9 or 10) compared to regions with earlier winter holidays (week 6, 7 or 8). Arnarson (2021) and Björk et al. (2021) demonstrate that these regions experienced markedly higher COVID-19 case rates during the first wave, likely exacerbated by the return of ski tourists from the Alps. For instance, Arnarson (2021) shows regions that had their winter holiday in week 9 reported almost twice as many COVID-19 cases in March 2020 compared to those with winter holiday in week 7.

Figure 1 below illustrates the timing of the winter holiday across European regions and highlights several notable aspects. Firstly, it seems to confirm the findings from Arnarson (2021) and Björk et al. (2021) as all regions in Belgium – the European country with the highest number of COVID-19 deaths per capita in spring 2020 – had their winter holiday in week 9. However, the figure also demonstrates that the timing of the winter holiday is not deterministic in terms of COVID-19 outcomes, as evidenced by Norway and Finland. Both countries have regions where the winter holiday occurs in week 9, yet, unlike Sweden, they experienced very few COVID-19 deaths during the pandemic.

**Figure 1.**
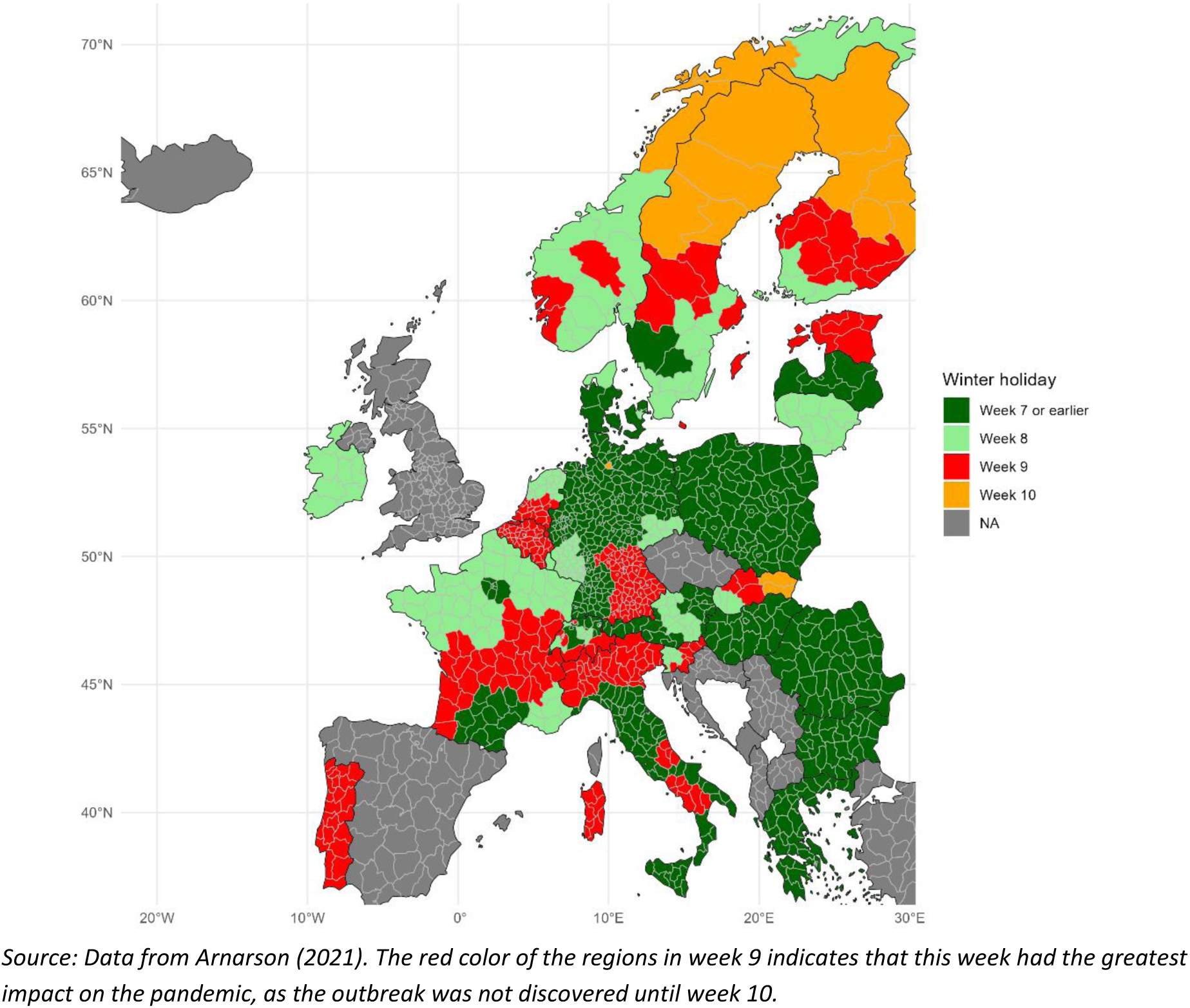
Timing of the winter holiday in European regions.

However, an important difference exists between these countries. According to Anders Tegnell’s recent book, approximately 1 million Swedes returned from international travel during the winter holidays, predominantly visiting the Alps during weeks 9 and 10. In contrast, Norwegians and Finns are less likely to travel to the Alps during these weeks, typically preferring vacations within their own countries, Tegnell (2023).

This behavior is corroborated by official flight data, as illustrated in Figure 2. Panel A displays the number of arrivals from the Alps by country and week in 2020, and Panel B highlights the deviations in these numbers from the 2019 average. The data reveal that Swedish arrivals from the Alps were approximately five times higher than those of Norwegians and Finns. This corresponds to data provided by Arnarson (2021) which shows that Swedes were more than twice as likely to spend a night in Austria during February 2020 compared to Norwegians and Finns.^8^

**Figure 2.**
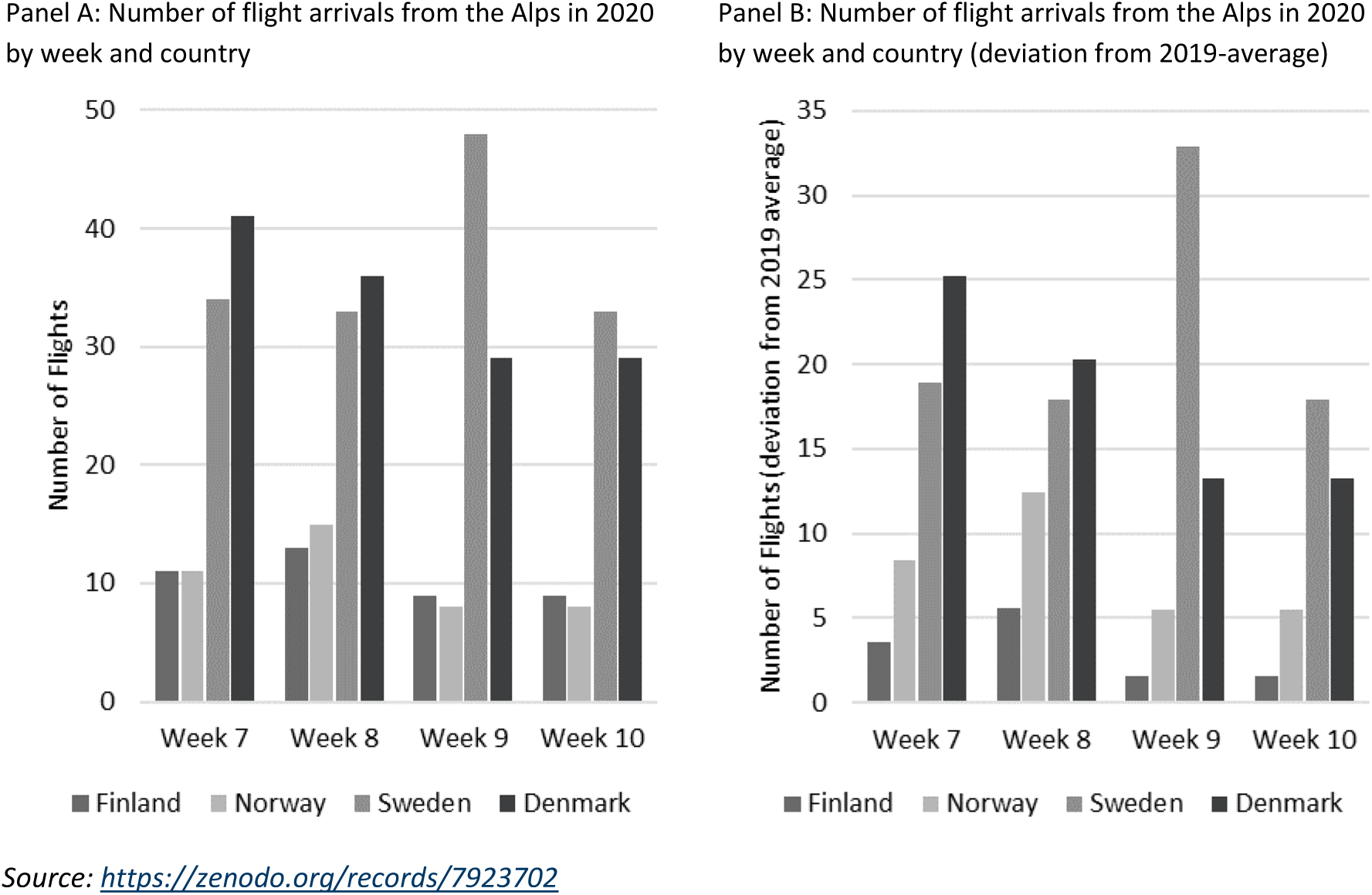
Skiing in the Alps.

Interestingly, while many Danes also traveled to the Alps during this period, Denmark’s COVID-19 death rate during the first wave was about double that of Norway and Finland, but only about one-fifth compared to Sweden. This indicates that while the timing of the winter holiday was a significant factor, it is far from being the sole determinant of the differences in COVID-19 outcomes.

The data presented in Figure 2 indicates that Swedes were more likely than Norwegians and Finns— and to a certain extent, Danes—to travel to the Alps during the weeks coinciding with an undisclosed COVID-19 outbreak. This was particularly true for week 9, during which Arnarson (2021) identified the most substantial impact of the virus.

## Results for Sweden

I begin by analyzing the impact of a Swedish lockdown in week 11 of 2020, utilizing a pre-intervention period starting from week 27 of 2016 and extending to a post-intervention period ending in week 26 of 2021, when vaccines were widely distributed to vulnerable residents. By selecting week 27 in 2016 as the commencement of the pre-intervention period, rather than week 1, I aim to avoid the influence of random peaks in mortality rates that could occur during the 2015/16 winter, which might otherwise introduce variability depending on whether they fall in 2015 or 2016. This approach ensures that the synthetic control remains unaffected by such random fluctuations. Based on these data boundaries the available donor pool consists of 36 countries.^9^

Figure 3 below displays the results of the SCM. Panel A shows the full pre- and post-intervention window of the analysis while Panel B focuses on the post-intervention period. Panel C shows the (cumulative) excess deaths for the post-intervention period based on the weekly differences between actual and synthetic Sweden. The donors are Netherland (37%), Canada (30%), Finland (23%), Denmark (9%), and Luxembourg (1%) (donor weights are available in Appendix C).

**Figure 3.**
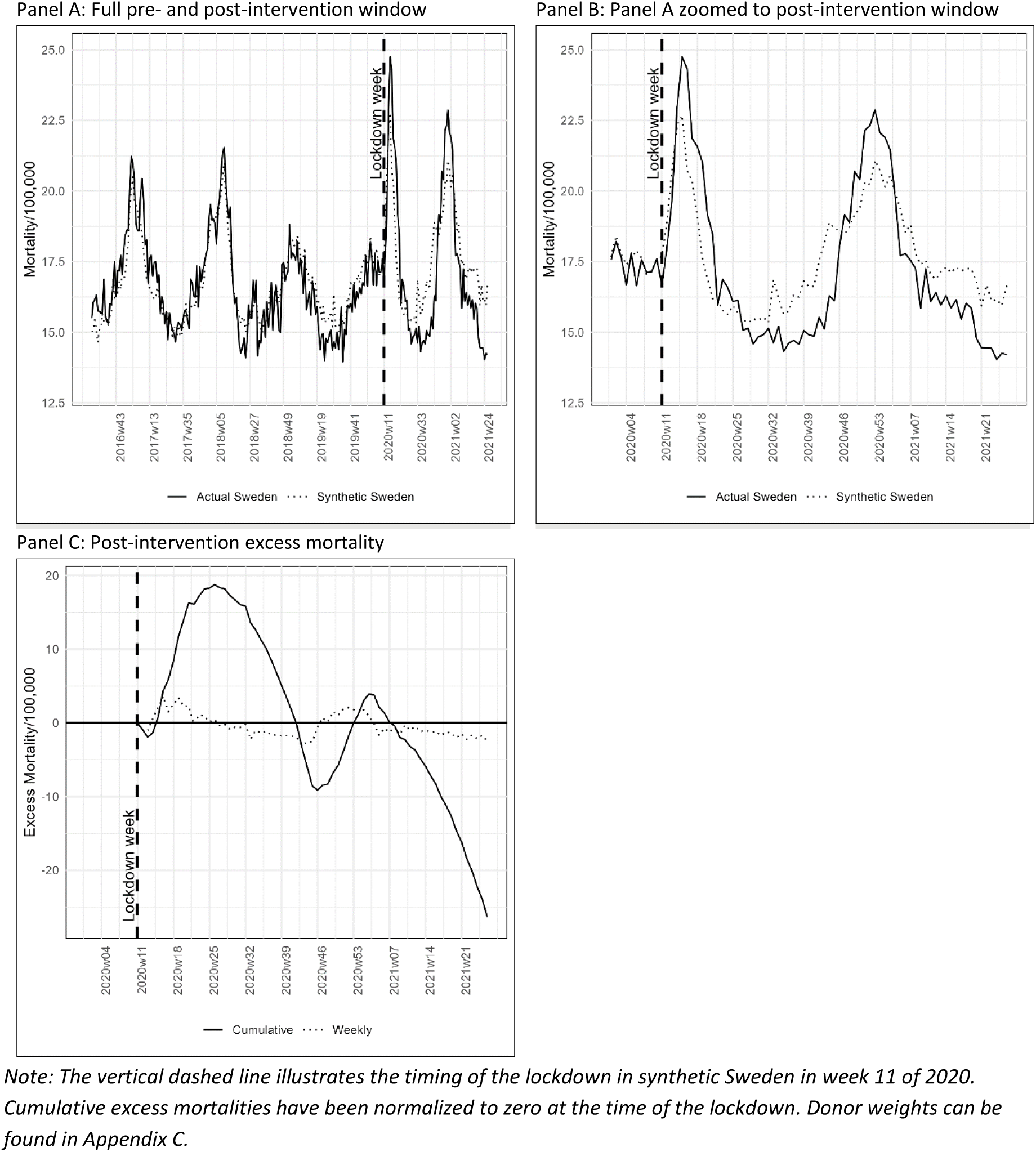
Results from the SCM for Sweden.

Figure 3 reveals some interesting findings. First, Panel A shows Synthetic Sweden’s ability to track Actual Sweden prior to lockdown. In general, Synthetic Sweden tracks Actual Sweden closely.

Second, Panel B shows that the mortality rate in Actual Sweden is higher compared to Synthetic Sweden during the first and second wave of the pandemic, but lower between and after the waves.

Third, Panel C shows that my results in the short run (the summer of 2020) are comparable to earlier findings. After the first wave (week 26 of 2020), the cumulative mortality in Actual Sweden is approximately 18 deaths per 100,000 higher than in Synthetic Sweden corresponding to approximately 1,900 avoided deaths in total. In comparison, both Conyon and Thomsen (2021) and Ege et al. (2023) estimate an effect of approximately 30 deaths per 100,000 (3,100 deaths in total), and Born et al. (2021) estimate that a lockdown would have reduced deaths by 38% corresponding to approximately 2,000 avoided deaths. In a Danish context, my results correspond to approximately 1,000 avoided deaths with a population of 5.8 million.

Fourth, Panel C also shows that the effect is reversed in the long run. At the end of the post-intervention period, the cumulative mortality in Actual Sweden is more than 25 deaths per 100,000 *lower* compared to Synthetic Sweden, corresponding to approximately 1,500 extra deaths with a Danish population.

### Robustness checks

The estimated short run effect of a counter factual Swedish lockdown is relatively small compared to the effects predicted ex ante by epidemiological studies. And in the longer run the effect is reversed so a lockdown causes more deaths. To examine the validity of the estimates I carry out a range of robustness checks.

#### Placebo test

The placebo test in the Synthetic Control Method (SCM) is a robustness check designed to assess the validity of the estimated treatment effect by comparing it to the distribution of estimated effects for untreated units. This test involves creating synthetic controls for units that were not exposed to the intervention and then evaluate whether these groups reveal a placebo treatment effect.

In practice, the placebo test is conducted by applying the SCM to each untreated unit in the donor pool, generating synthetic controls as if these units had received the intervention (no lockdown). The distribution of these placebo effects is then compared to the actual estimated effect of the intervention on the treated unit. If the observed treatment effect is substantially larger than the placebo effects, it provides stronger evidence that the estimated impact is not driven by chance or unobserved confounders. According to Abadie et al. (2010) and Abadie et al. (2015) the share of control units which exhibit outcomes similar to the treated unit is akin to a statistical p-value.

The results from the placebo test are illustrated in Figure 4 below. Since there are 36 donors the results are akin to statistically significant at the 5% significance level if at most two of the placebos exceeds the estimated effect for Sweden. This is clearly not the case. In all weeks following the lockdown there are several placebos experience higher cumulative mortality, so the results for Sweden cannot be interpreted as significant.

**Figure 4.**
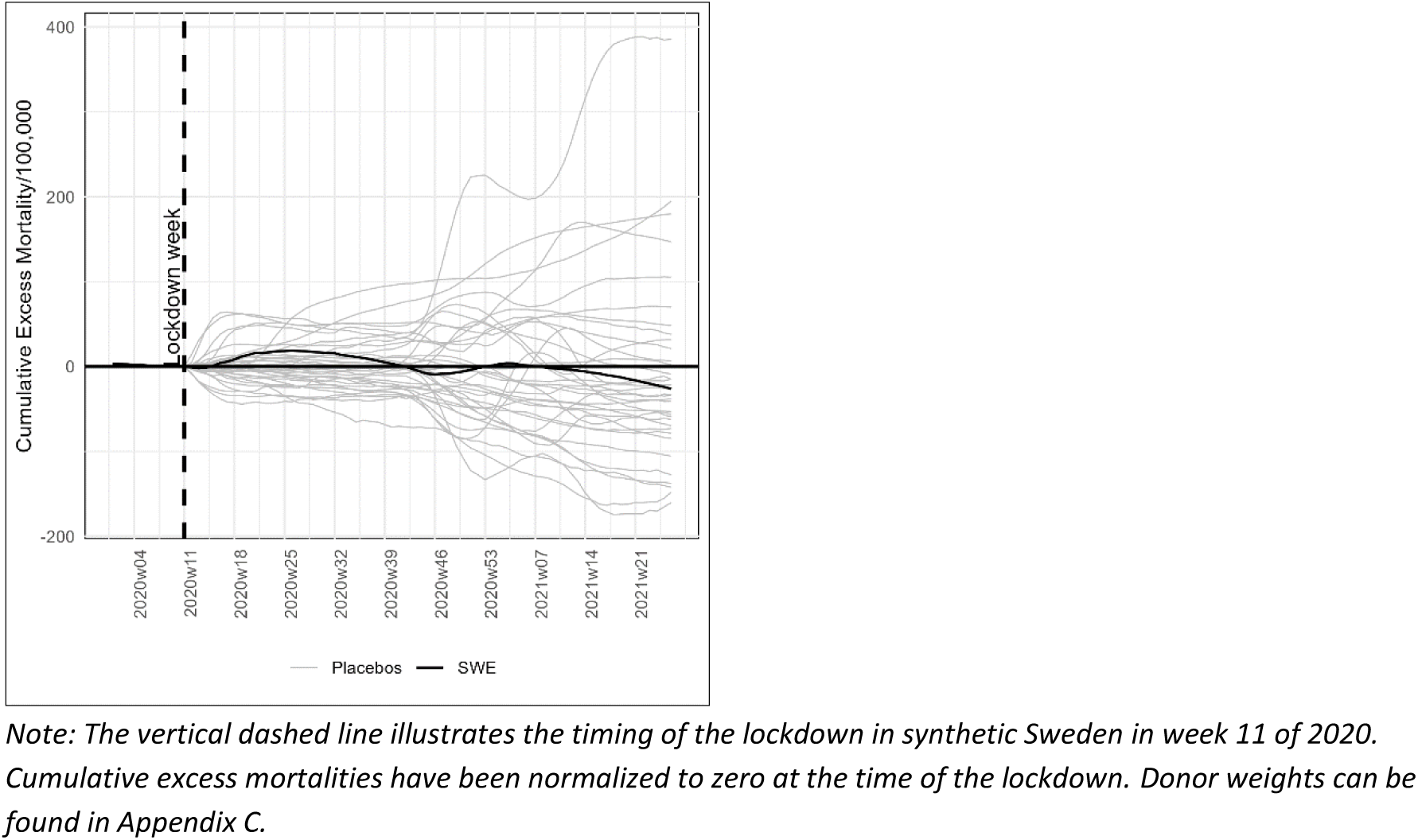
Placebo test for Sweden.

The results of my placebo test are similar to the results achieved by Ege et al. (2023). Ege et al. run a placebo test which show that 4-5 out of the 29 donor countries (14% - 17%) exhibit the same or higher excess mortality by the end of the study period.^10^ Based on the results of their placebo test Ege et al. conclude that their “estimated impact of not implementing a mandatory lockdown in Sweden is not likely to be observed by chance, and is clearly driven by the treatment in question.” But this interpretation conflicts with the results of their placebo test which show that the results in Ege et al. are not statistically significant.

#### Extending the pre-intervention window to 2011 or 2007

To test how much my results are affected by my choice to start the pre-intervention window in Week 27 of 2016, I run two supplementary analyses, where I set the start to week 27 of 2011 and week 27 of 2007 reducing the donor pool to 31 and 26 countries respectively.^11^ The results are shown in Figure 5 below.

**Figure 5.**
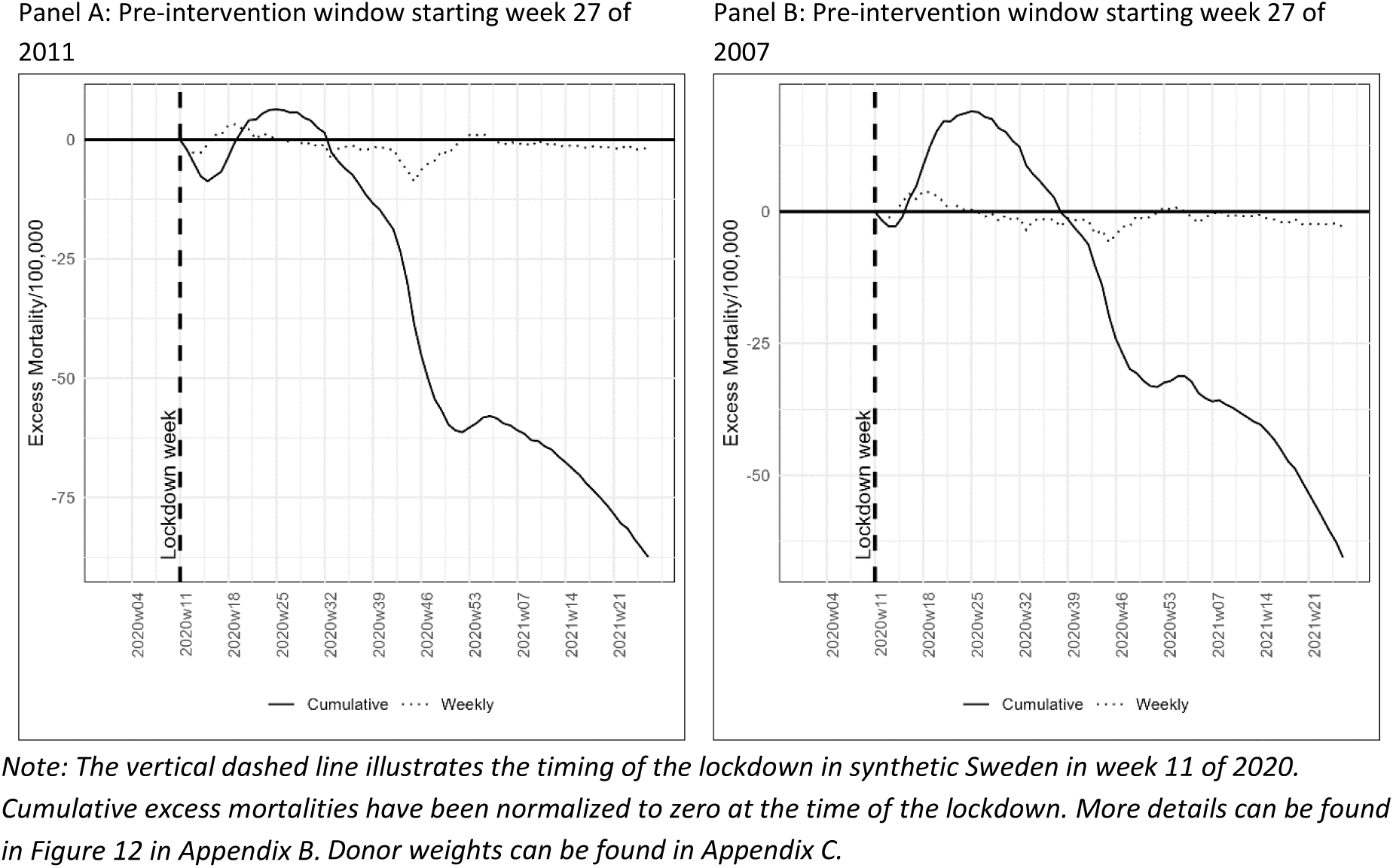
Estimated effect of lockdown when Synthetic Sweden is based on a longer pre-intervention window.

The figure illustrates some sensitivity to changes in the pre-intervention window. If the pre-intervention window starts in 2007 (Panel B), the results are similar to my main model in the short run, but find a larger negative effect of lockdowns in the longer run. If the pre-intervention window is set to start in 2011 (Panel A), I find virtually no effect in the short run and a large negative effect in the long run).

As with the main specification, none of the results turns out as significant in the placebo test (see Figure 12 in appendix B).

#### Extending post-intervention window to 2023

In the main analysis the treatment runs to week 26 of 2021, where vaccines were widely distributed. However, limiting the analysis to the middle of 2021 may yield a bias in the evaluation of the long run effects of lockdowns. For example, Mulligan and Arnott (2022) show that “From April 2020 through at least the end of 2021, Americans died from non-Covid causes at an average annual rate of 97,000 in excess of previous trends.” Hence, a longer post-intervention period may yield a better evaluation of the long run consequences of lockdowns.

In Figure 6 below I show the results when the post-intervention period is extended to week 26 of 2023. Panel A focuses on the post-intervention period (the pre-intervention period is identical to the main analysis illustrated in Figure 3, panel A), and panel B shows the results from the placebo test.

**Figure 6.**
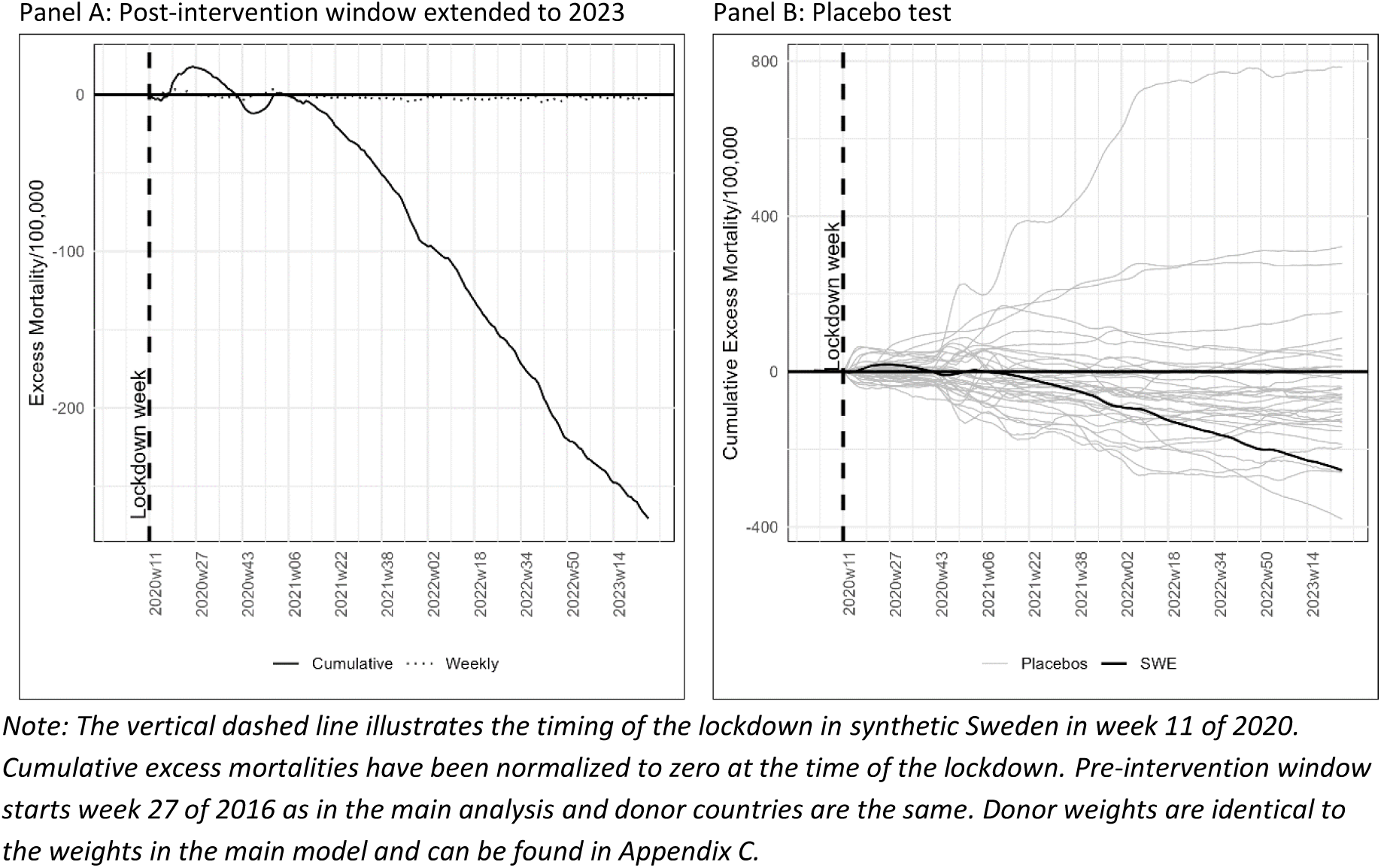
Results when post-intervention window is expanded to week 26 of 2023.

The results show that the tendency we have seen in my earlier results, where Sweden performs better in the longer run, continues after week 26 of 2021. Panel A in Figure 6 shows that in week 26 of 2023 – the end of the analyzed period – I estimate that a lockdown would have caused more than 250 extra deaths per 100,000 corresponding to more than 25,000 extra deaths in Sweden or 15,000 extra deaths in a population the size of Denmark’s. Panel B shows that this estimate is significant at the 5%-level, as just one other country (Israel) out of the 36 placebos (2,8%) estimate lower excess deaths by week 26 of 2023. Two other countries (Hungary and Iceland) have estimates very close to that of Sweden (-259 and -255 respectively, while the estimate for Sweden is -260), so one should be careful to draw too strong conclusions based on the result.

On possible explanation for the large estimated negative effect of a lockdown is that Sweden was superior in rolling out vaccines – a variable I cannot include in my analysis, as it happened after the pre-intervention period ends, and which may in itself be affected by the choice to lock down.^12^ However, there is no apparent difference between the rollout of vaccines in Sweden and the donor countries making an omitted variable bias with regards to vaccines less likely.^13^

## Results regarding the timing of the winter holiday

I now turn to explore the impact of the timing of the winter holiday and the decision not to implement lockdown measures. It is worth noting that I am not interested in the effect of the timing of the winter holiday per se. Rather, I use the timing of the winter holiday as a proxy for the extent/spread of COVID-19 in societies before lockdown – a variable which is unobservable due to limited testing.

To explore the impact of the timing of the winter holiday, I divided Sweden into four hypothetical countries, “Sweden w7”, “Sweden w8”, “Sweden w9”, “Sweden w10” based on the timing of the winter holiday in the Swedish regions (län). Table 1 shows the basic statics for the four hypothetical countries. For comparison, the population of Denmark, Norway, and Finland in 2020 was 5.8 million, 5.5 million, and 5.5 million respectively. Hence, three of the four hypothetical countries are slightly smaller than their Scandinavian neighbors but larger than e.g. the Baltic countries.

**Table 1.** Basic statistics for hypothetical countries.

| Hypothetical country | Population 2020 | Cumulative COVID-19 mortality per 100,000<br>by week 26, 2020 |
| --- | --- | --- |
| Sweden w7 | 2.100.000 | 44 |
| Sweden w8 | 3.790.000 | 35 |
| Sweden w9 | 3.590.000 | 79 |
| Sweden w10 | 900.000 | 30 |
| Sweden | 10.380.000 | 52 |

*Source: SCB Statistics and winter holiday data from Arnarson (2021)*

All four hypothetical countries maintained the same COVID-19 policies, notably abstaining from lockdowns in spring 2020. If the decision against lockdowns were an important cause of excess mortality, we would expect similar excess mortalities in all four hypothetical countries. Contrary, if lockdowns had little effect and the differences in pandemic outcomes were driven by unobserved extent/spread of COVID-19 in societies before lockdown, we would expect to find large differences in the estimated effect with a large effect in Sweden w9 and possibly Sweden w10 and smaller or no effects in Sweden w7 and Sweden w8.

Since I don’t have data for hospital beds, urban population etc. for the four hypothetical countries, I apply the data for Sweden to all four hypothetical countries. As in my main analysis, the pre-intervention period starts in week 27 of 2016 and the post-intervention analysis ends in week 26 of 2021.

Figure 7 below presents the outcomes of the analysis. It shows that the estimated effect of a Swedish lockdown varies greatly depending on the timing of the winter holiday. In the short run, the estimated effect is by far largest in Sweden w9, whereas the estimated effect is very small in Sweden w8 and Sweden w10. In Sweden w7, the effect it similar to that of Sweden as a whole.

**Figure 7.**
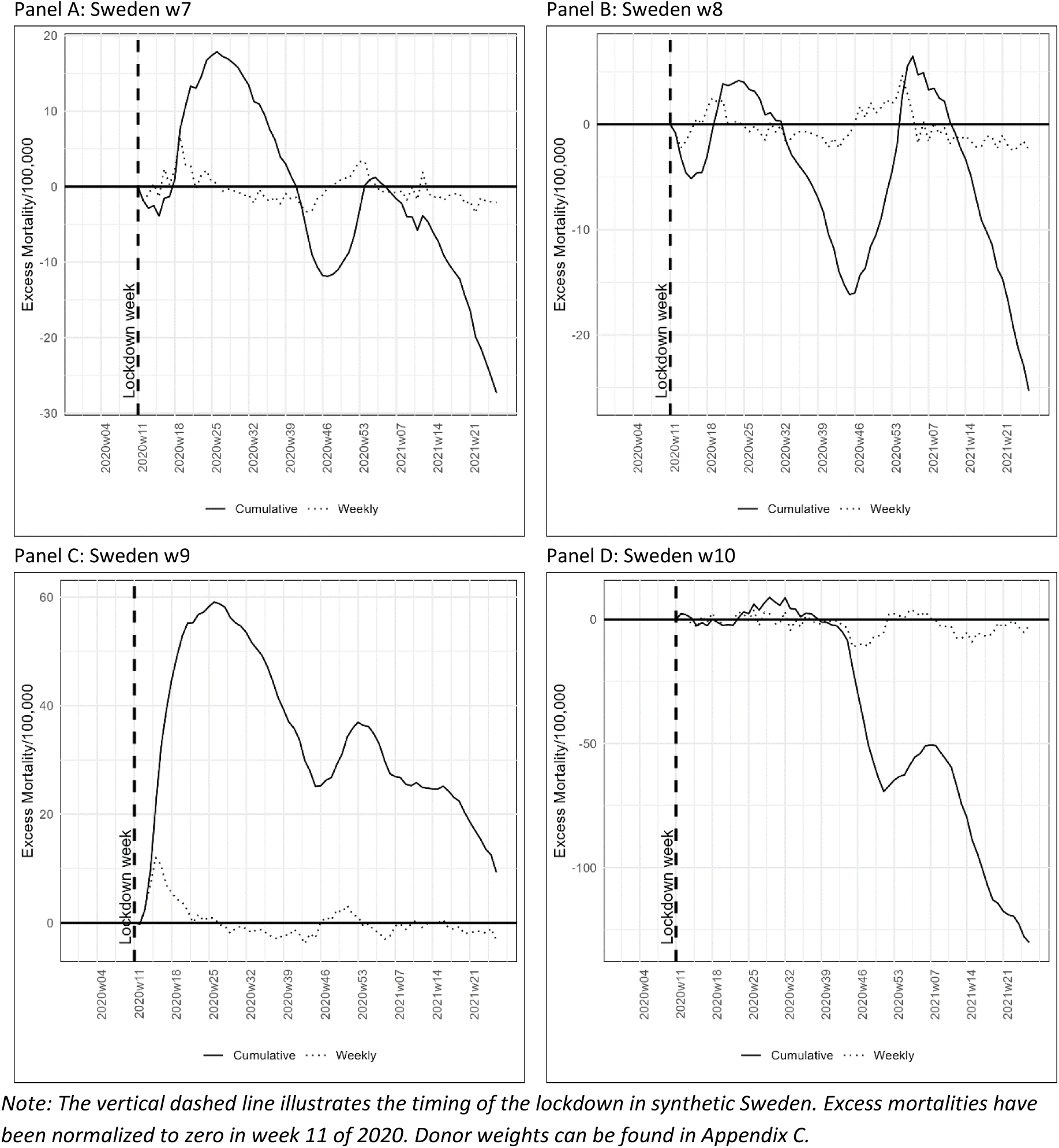
Post-intervention excess mortalities for hypothetical countries.

Focusing on the end of the analyzed period (week 26 of 2021) I find a large negative effect of a lockdown in Sweden w7, Sweden w8, and especially Sweden w10. In Sweden w9, I estimate a positive effect of approximately 10 avoided deaths per 100,000.

Figure 8 below show the results of the placebo test for the four hypothetical countries. The placebo test show that there is a statistically significant difference between Sweden w9 and the synthetic control in the short run, but not in the longer run.

**Figure 8.**
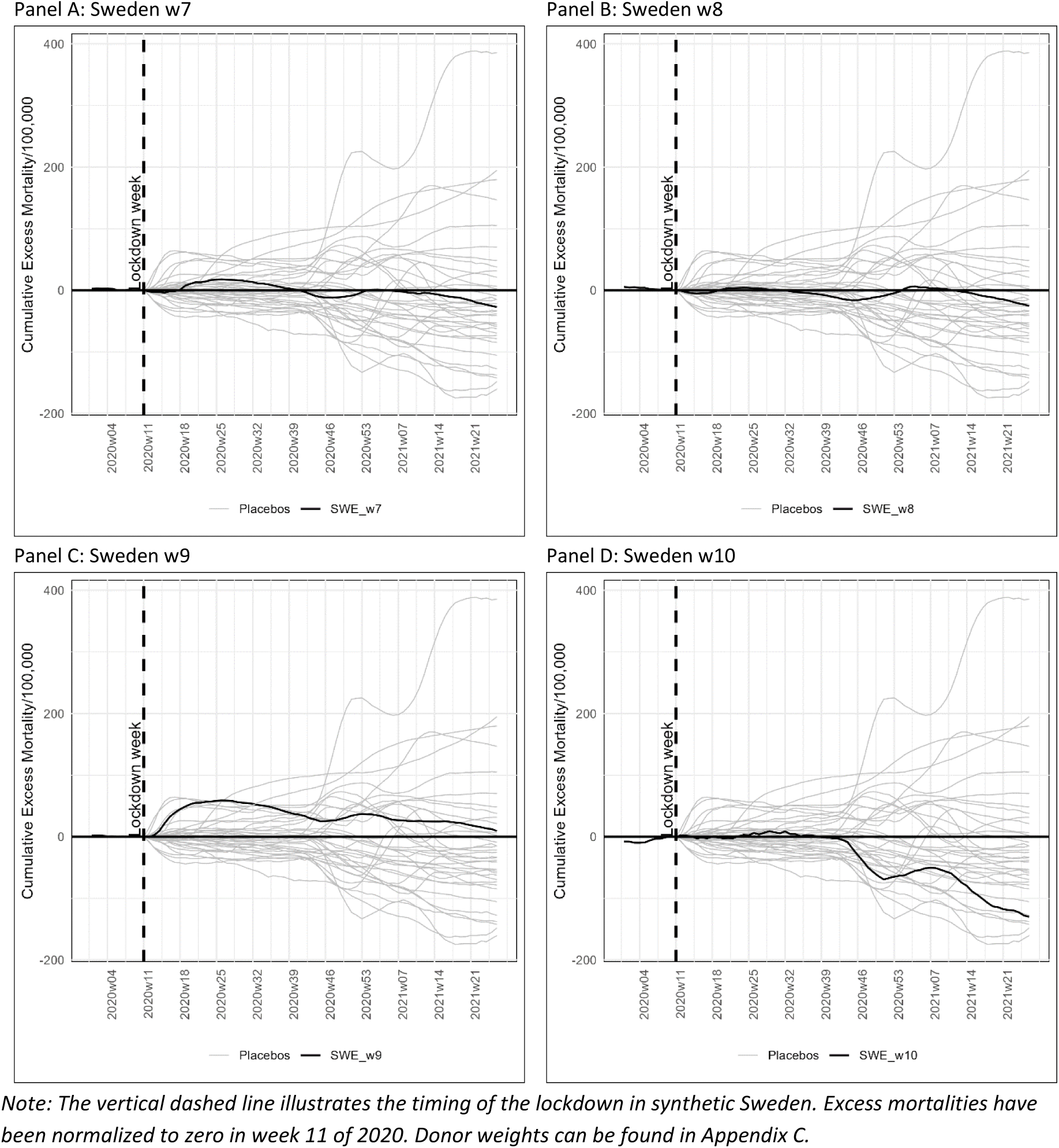
Placebo tests for hypothetical countries.

However, it is unlikely to be caused by Sweden’s decision not to lock down, as we do not see a similar effect in the other hypothetical countries. Instead, the apparent significant results are likely to be driven by the unfortunate timing of the winter holiday in Sweden w9 (as shown by Arnarson (2021) and Björk et al. (2021)) and possibly other unobservable variables.

## The limits of the SCM in evaluating the effect of a counter factual lockdown in Sweden

Earlier studies of the effect of a counter factual lockdown in Sweden using COVID-19 data such as positive COVID-19 cases or deaths are constrained by very short pre-intervention windows as countries in the donor pool typically locked down within weeks after the first case. The very short pre-intervention windows severely limit the SCM, as “it is of crucial importance to collect information on the affected unit and the donor pool for a large pre-intervention window” (Abadie (2021)).

Some studies, including the present study, try to handle the problem with very short pre-intervention windows by using historical mortality rates rather than COVID-19 measures. However, this approach carries its own problems, as mortality rates in the past may not be a precise predictor for how countries were impacted by the COVID-19 in the first weeks of the pandemic. The results in Arnarson (2021) and Björk et al. (2021) are concrete examples as they show that the timing of the winter holiday had a notable effect on the spread of COVID-19 in Europe, whereas the timing of the winter holiday is unlikely to play a notable effect on mortality rates pre-pandemic.

The large discrepancy in the effect estimates across hypothetical countries with varying winter holiday timings suggest that differences in excess mortality are less attributable to the absence of a lockdown policy and more to unobserved variables, such as the extent and spread of COVID-19 in societies prior to lockdown, which I approximate using the timing of winter holidays. These unobserved variables can introduce substantial biases into the SCM because seemingly minor choices may cause significant shifts in donor weights, potentially leading to large differences in mortality outcomes in the synthetic controls.

For instance, the large discrepancy in the effect estimates across the hypothetical countries are largely driven by shifts in donor weights. Bulgaria—a country that experienced very high excess mortality during the winter of 2020/21—has zero weight in the synthetic controls for Sweden w7, Sweden w8, and Sweden w9, but a significant weight of 24% in the synthetic control for Sweden w10. As a result, the high excess mortality estimated for Sweden w10 is largely due to Bulgaria’s inclusion in the synthetic control.

This is illustrated in Figure 9 below, which shows mortality rates in the synthetic controls for the four hypothetical countries. The figure indicates that while the synthetic controls are somewhat similar during the first pandemic wave, the synthetic control for Sweden w10 diverges markedly in the longer run.

**Figure 9.**
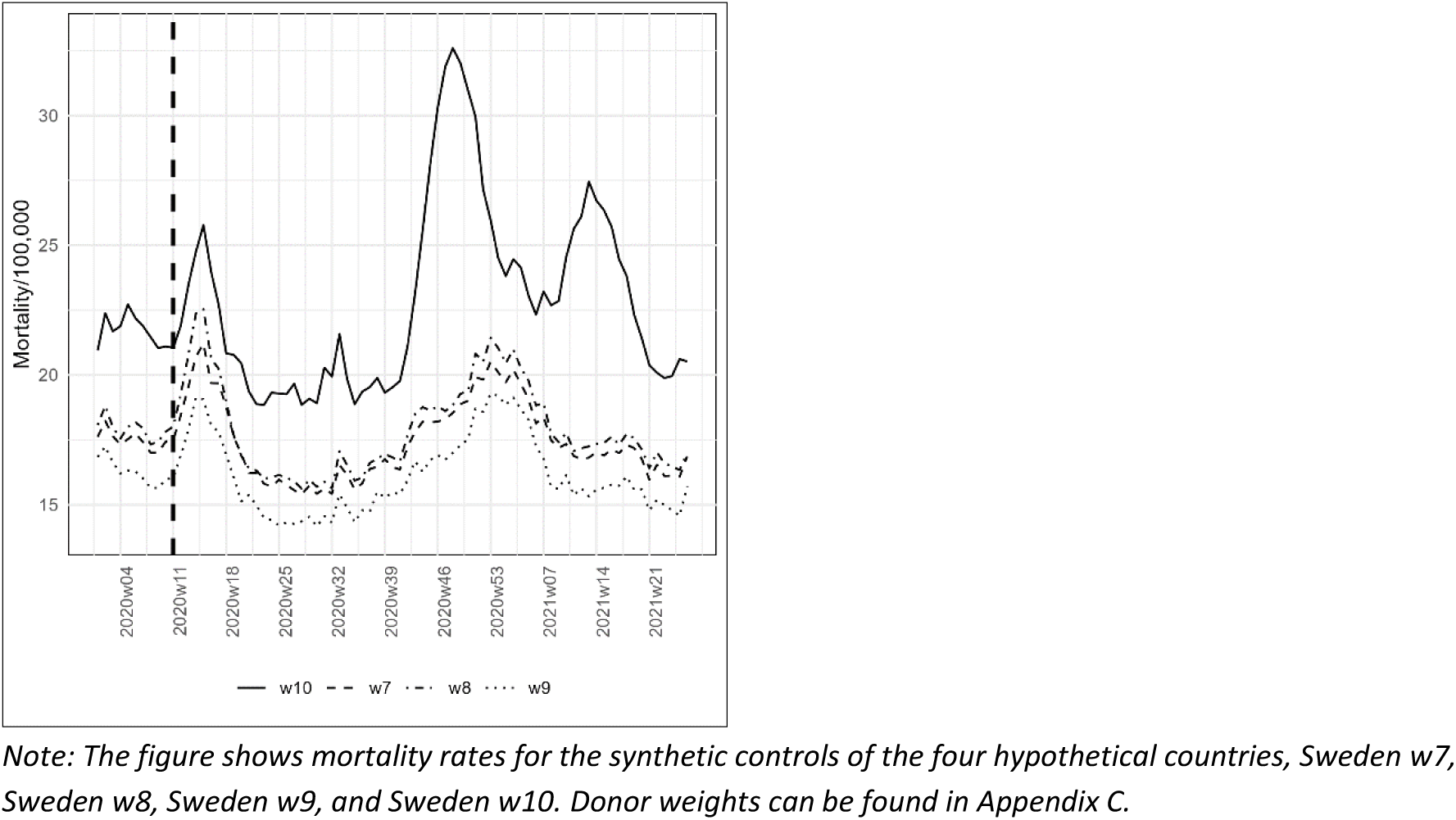
Morality rates for the synthetic controls of the four hypothetical countries.

These insights indicate that the effect of lockdowns found in other SCM studies using Sweden as a control are – at least partially – driven by unobserved variables.

### Causality problems are minor

Another potential limitation is the potential causality problem. If increasing infection rates lead governments to introduce lockdown policies, and declining infection rates subsequently lead them to ease lockdowns, the estimated association between policy stringency and mortality may be biased. It is essential for the SCM that the treatment is the only important factor that differs between treatment and control group in the post-treatment period. This may not be satisfied if there is a great degree of reverse causality and may explain why I do not find any effect of lockdowns.

However, for a number of reasons I believe that this potential bias is likely to be very small.

In the beginning of the pandemic there was very little information available to governments because of very limited testing capacities. This means that governments could not react based on information about infections. Rather, Sebhatu et al. (2020) show that government policies were strongly driven by the policies initiated in neighboring countries rather than by the severity of the pandemic in their own countries. In short, Sebhatu et al. show that it is not the severity of the pandemic that drives the adoption of lockdowns, but rather the propensity to copy policies initiated by neighboring countries.^14^ Similar results are found by Engler et al. (2021) and Mistur et al. (2023), while Bjørnskov and Voigt (2022) show that the use of lockdown was influenced by the additional emergency powers granted by the constitution.

These insights are illustrated in Figure 10 below which shows the correlation between the spread of infection on 20 March 2020 (proxied by the daily death toll three weeks later) and the degree of lockdown. The figure shows that there was a positive correlation, so that countries that had a high spread of infection also shut down more strictly. But the correlation is very small and by no means statistically significant.

**Figure 10.**
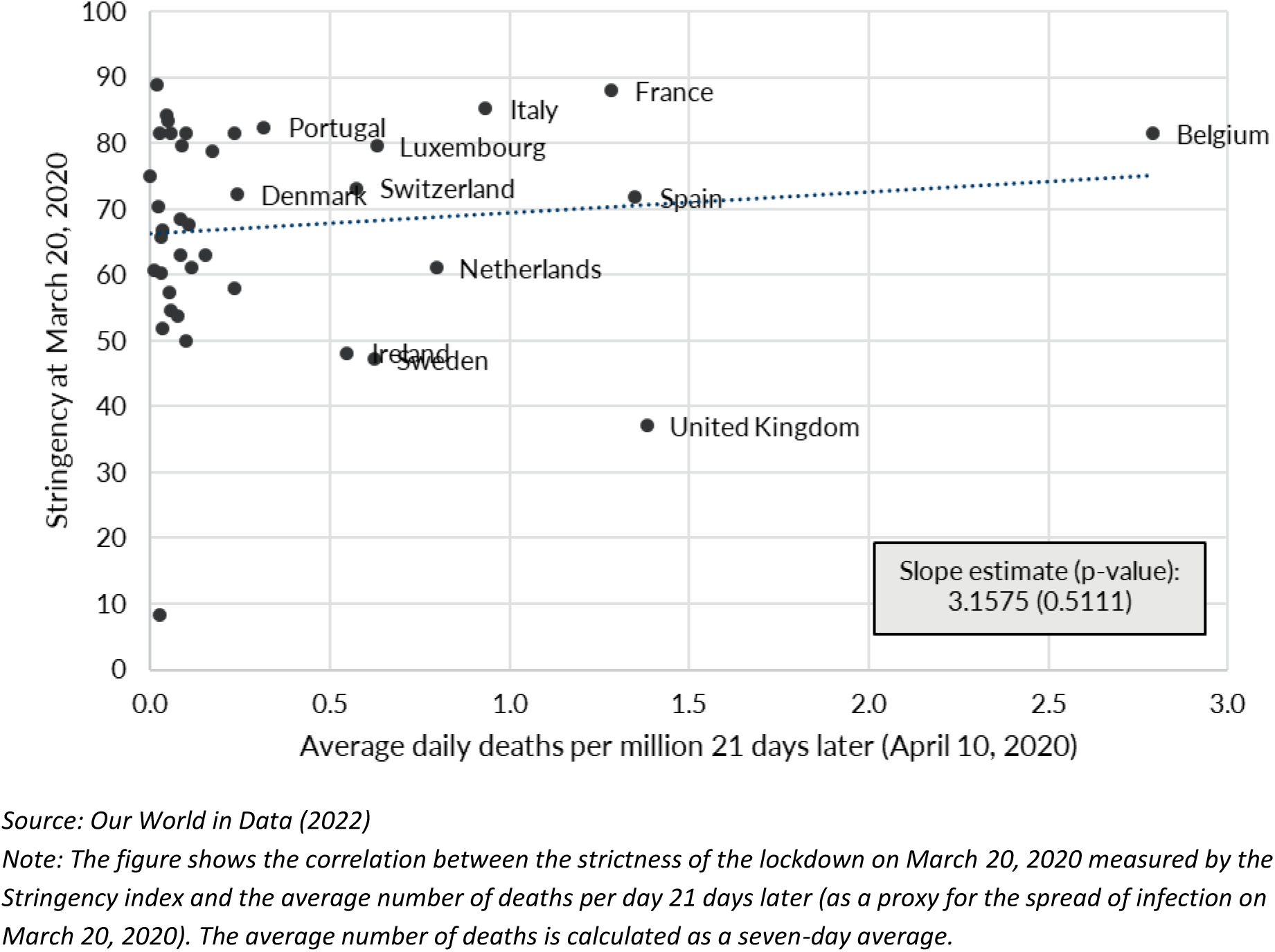
There was no correlation between the spread of infection and the degree of lockdown in the spring of 2020.

According to Sebhatu et al. (2020), this kind of imitation is common among decision-makers when they are unsure about the effects of a decision. By doing what everyone else does, decision-makers can avoid being criticized for being too slow or not responding at all (which Sweden received a lot of criticism for). In addition, politicians can adopt rules that are similar to those of other countries to signal that they are part of the “normal”.

## Conclusion

Contrary to Denmark, Finland, and Norway, Sweden experienced significant COVID-19 mortality in the spring of 2020 and did not enforce a mandatory lockdown. These contrasts led many to jump to the conclusion that the absent lockdown in Sweden caused a larger than necessary spread of the virus and consequent excess mortality. Numerous studies using the SCM supported this initial assumption. However, as I argue here, these studies may possess critical flaws, notably the short pre-intervention windows and the winter holiday timing combined with vacation patterns of Swedes, particularly their travels to the Alps during the winter holidays where a significant, initially undisclosed outbreaks occurred, in week 9 of 2020.

These flaws have mostly been neglected although the results of many of these studies should have raised major concerns. For instance, Ege et al. (2023) find that excess deaths in “Sweden and synthetic Sweden start to diverge immediately” after the lockdown despite the widely recognized epidemiological fact that the period from infection to death usually spans at least two weeks (see Appendix D).

In the present study, I first address two potentially critical flaws in the existing literature based on the Synthetic Control Method (SCM) which typically uses a very short pre-intervention period (sometimes limited to a few weeks) and only examines the short run effect of lockdown. Specifically, I extend the pre-intervention window substantially using mortality rates from week 27 of 2016 to week 11 of 2020. This pre-intervention period is far longer than any existing study. I also extend the post-intervention window to week 26 of 2021, where vaccines were distributed to the most vulnerable citizens, effectively ending the severe part of the pandemic. The longer post-intervention period allows a more in-depth analysis of potential trade-offs between short run benefits and long run costs of lockdowns.

I find that a counter factual lockdown would have reduced mortality rates by approximately 18 deaths per 100,000 in the short run (week 26 of 2020), but *increased* mortality by more than 25 deaths per 100,000 in the longer run (week 26 of 2021). However, these results are statistically insignificant. When I extend the post-intervention period to week 26 of 2023 I estimate that a counterfactual lockdown in Sweden would have caused more than 250 extra deaths per 100,000 corresponding to more than 25,000 extra deaths in Sweden. This estimate is (borderline) significant, but I warn about drawing too strong conclusions based on the result, as minor changes in e.g. the pre-intervention window are likely to affect the result.

I then examine the impact of the timing of the winter holiday in Sweden as a proxy for the extent/spread of COVID-19 in societies before lockdown – a variable which is unobservable due to limited testing. I find large discrepancies in the effect estimates across regions with different timing of their winter holiday suggesting that differences in excess mortality are less a result of the no-lockdown policy and more a result of unobserved variables such as the extent/spread of COVID-19 before lockdown. This result indicate that the effect of lockdowns found in other studies using Sweden as a control are at least partially driven by unobserved variables.

My findings suggest that a counter factual lockdown in Sweden would not have yielded a substantial benefit and may even have had an adverse long-run effect on excess mortality. However, it is crucial to approach these conclusions with caution as other factors could have significantly influenced the observed outcomes. Consequently, one of the critical takeaways is the need for caution when using the SCM to evaluate the effects of policy interventions such as lockdowns. While SCM is a powerful and intuitive tool, it may inadvertently fail to account for critical omitted variables, potentially skewing the interpretation of the true impact of lockdown measures.

Unfortunately, many SCM studies tend to focus on outliers with notably high COVID-19 mortality rates during the first wave. Herby et al. (2023) report that at least seven out of eleven eligible SCM studies targeted regions that experienced substantial deaths early in the pandemic.^15^ This focus likely introduces selection bias, which could lead to misleading conclusions about the general effectiveness of interventions. In contrast, Mader and Rüttenauer (2022) utilized the Generalized Synthetic Control Method (GSCM) across a broader dataset of 169 countries, effectively minimizing such biases. Their findings reveal no significant or consistent mitigating effects of non-pharmaceutical interventions (NPIs) on COVID-19-related deaths per capita. This disparity underscores the importance of employing diverse methodologies and comprehensive data sets to avoid skewed interpretations and better understand the true impact of public health measures.

## Data Availability

All data produced are available online at https://github.com/JonasHerby/Dont-Jump-to-Faulty-Conclusions

## Appendixes

## Appendix A Using cumulative excess mortalities in Ege et al. (2023)

The analysis in Ege et al. starts in week 44 of 2019, where the cumulative excess mortality equals the excess mortality for that week, effectively setting week 43 as the zero point for accumulation. Figure 11 below presents the results from Ege et al., alongside an identical SCM where the only change is setting week 44 as the zero point for accumulation. This adjustment is made by subtracting the value in week 44 from all subsequent weeks by country.

**Figure 11.**
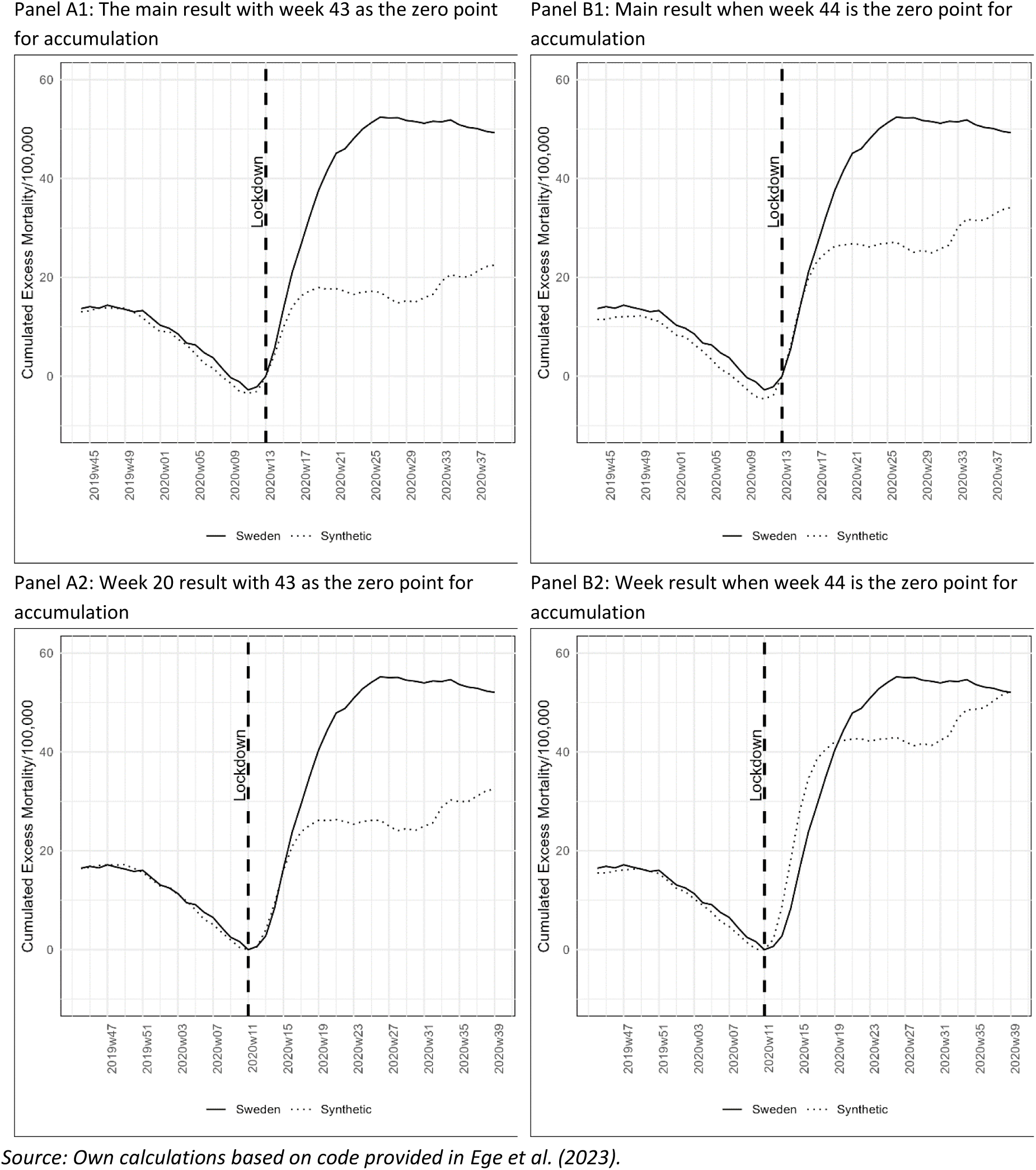
Selecting Other Weeks as Zero Points for Cumulative Excess Mortality in Ege et al. (2023)

The top panels of Figure 11 illustrates the result when the lockdown week is set to week 13 of 2020 (the main specification in Ege et al.), while the bottom panels row shows the result when the lockdown week is set to week 11 of 2020 (the secondary specification). The first column, panel A, presents the original results from Ege et al., and the second column, Panel B, shows the results when the zero point for accumulation is shifted by one week.

Figure 11 demonstrates that adjusting the zero point for accumulation by one week significantly alters the results markedly. In the main specification (Panel A), the estimated effect of the lockdown is almost halved, and in the secondary specification (Panel B), the effect of the lockdowns disappears entirely.

## Appendix B Extra figures for pre-intervention window extended to 2011 and 2007

Figure 12 below shows the full period and placebo tests for both alternative pre-intervention window

**Figure 12.**
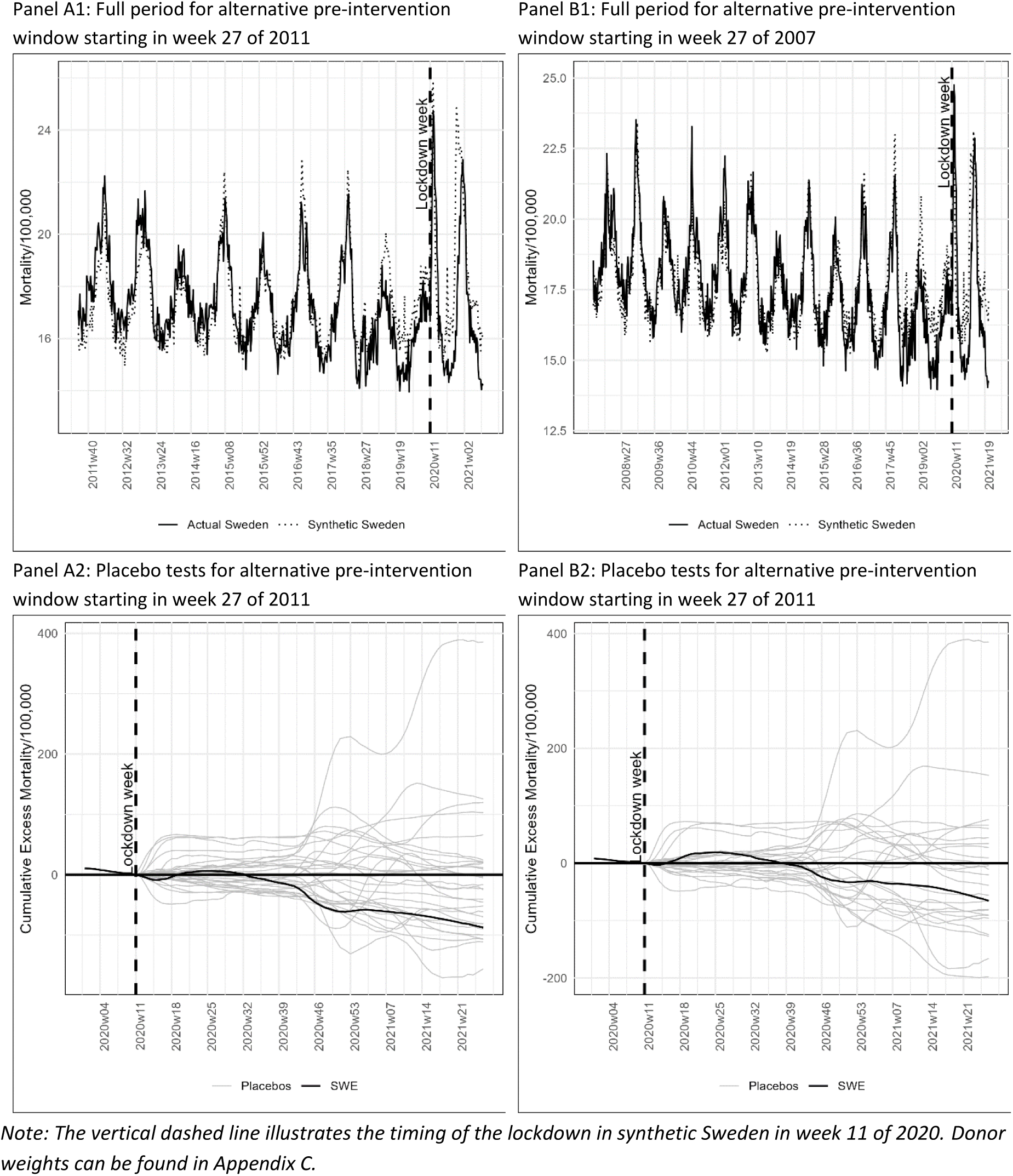
Full pre-intervention periods and placebo tests for alternative pre-intervention windows.

When the pre-intervention window starts in week 27 of 2011, the donor pool comprises 31 countries. When the pre-intervention window starts in week 27 of 2007, the donor pool comprises 27 countries. This means that the results for Sweden are statistically significant if at most two of the placebo tests find larger effects. As seen in row two, this is not the case for any of the alternative pre-intervention windows.

## Appendix C Donor weights from the SCM model

**Table 2.** Donor weights from the SCM model for the counterfactual synthetics.

| Donor country | Sweden<br>(2016) | Sweden<br>(2011) | Sweden<br>(2007) | Sweden w7<br>(2016) | Sweden w8<br>(2016) | Sweden w9<br>(2016) | Sweden w10<br>(2016) |
| --- | --- | --- | --- | --- | --- | --- | --- |
| DNK | 9,0% | 0,2% | 52,4% | 17,2% | 25,4% | 33,2% | 21,4% |
| FIN | 22,8% | 29,7% | 0,0% | 24,6% | 20,4% | 0,0% | 23,3% |
| CAN | 29,7% | 0,6% | NA | 33,2% | 20,1% | 32,5% | 0,0% |
| NLD | 37,3% | 0,7% | 0,0% | 22,1% | 33,9% | 17,7% | 0,0% |
| BEL | 0,0% | 31,3% | 18,2% | 0,0% | 0,0% | 0,0% | 22,4% |
| CHE | 0,0% | 34,2% | 20,3% | 0,0% | 0,0% | 0,0% | 0,0% |
| BGR | 0,0% | 0,0% | 0,0% | 0,0% | 0,0% | 0,0% | 24,0% |
| LUX | 1,1% | 0,0% | 5,9% | 2,2% | 0,0% | 1,8% | 8,9% |
| ISR | 0,0% | 0,0% | 0,0% | 0,0% | 0,0% | 13,0% | 0,0% |
| LVA | 0,0% | 0,0% | 3,1% | 0,0% | 0,0% | 0,0% | 0,0% |
| ISL | 0,0% | 0,0% | 0,0% | 0,0% | 0,0% | 1,5% | 0,0% |
| ESP | 0,0% | 1,3% | 0,0% | 0,0% | 0,0% | 0,0% | 0,0% |
| Others | 0,1% | 2,0% | 0,1% | 0,7% | 0,2% | 0,3% | 0,0% |

*Note: Only countries with a weight of at least 1.0% in one of the models are included in the table. The year in parentheses indicate the year the pre-intervention window starts.*

## Appendix D The time from infection to death

There is considerable variability in the time from infection to death. Virtually no-one dies within the first week of infection, but thereafter the probabilities climb quite steeply, cf. Figure 13 from Wood (2022).^16^ On average, it takes 26 days from infection to death, and 29% die within the third week after infection (day 15 to 21) while 26% die within the fourth week after infection (day 22 to 28).

**Figure 13.**
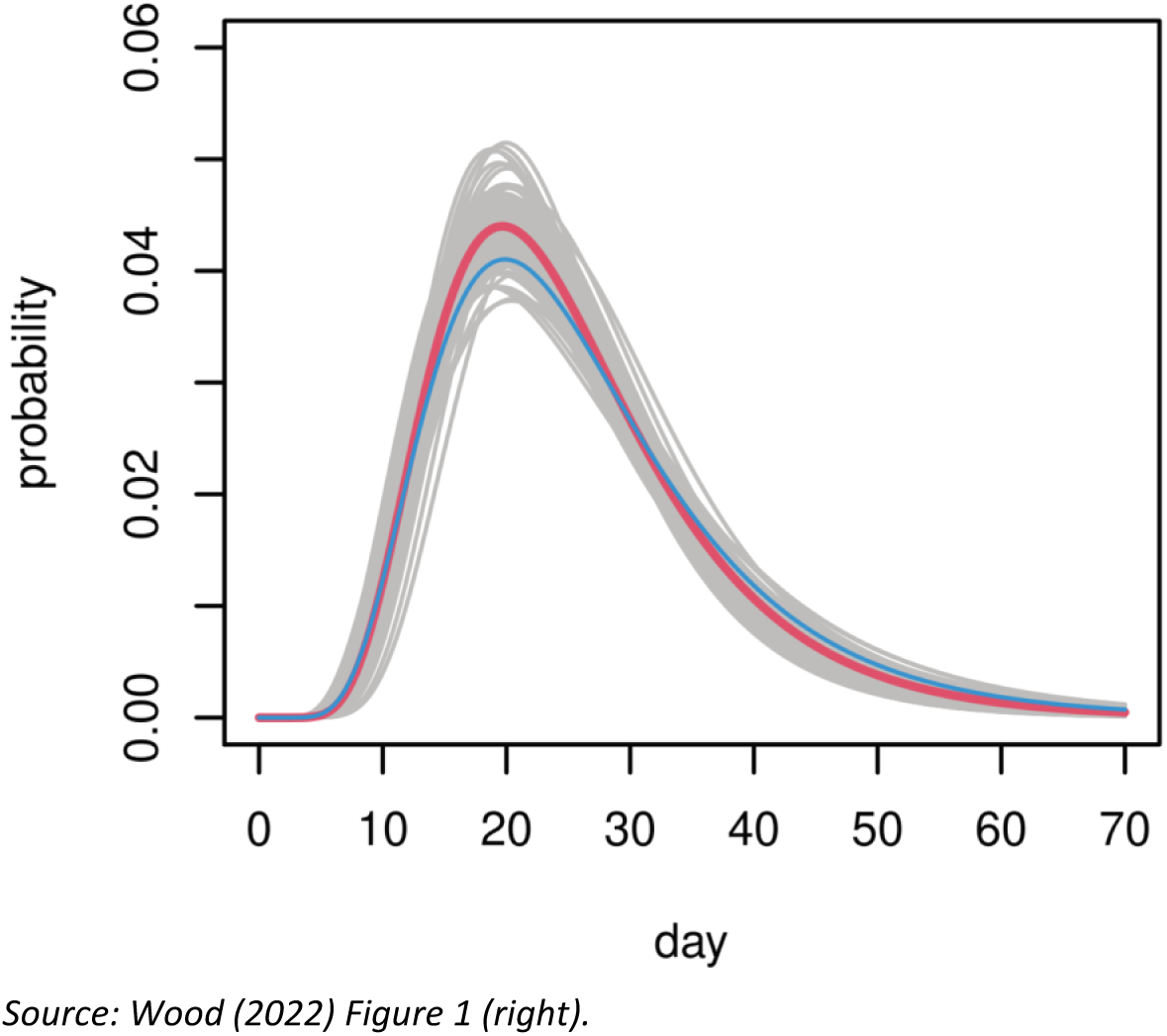
Fatal disease duration distribution from infection to death.

Because the duration between infection and death is probabilistic rather than deterministic, there will be a delay before the possible effect of a lockdown is visible in mortality data.

To illustrate this point, Figure 14 below shows the development in COVID-19-deaths from a simple SEIR-model which is deterministic and assumes that 0.5% of all infected dies every day compared to the same model where the risk of death is probabilistic.^17^ Figure 14 illustrates that in this case, where the reproduction rate is 2,584 before lockdown and 0,704 after, the effect of the lockdown is hardly visible in the cumulative number of deaths before day 14-16 after the (effective) lockdown.

**Figure 14.**
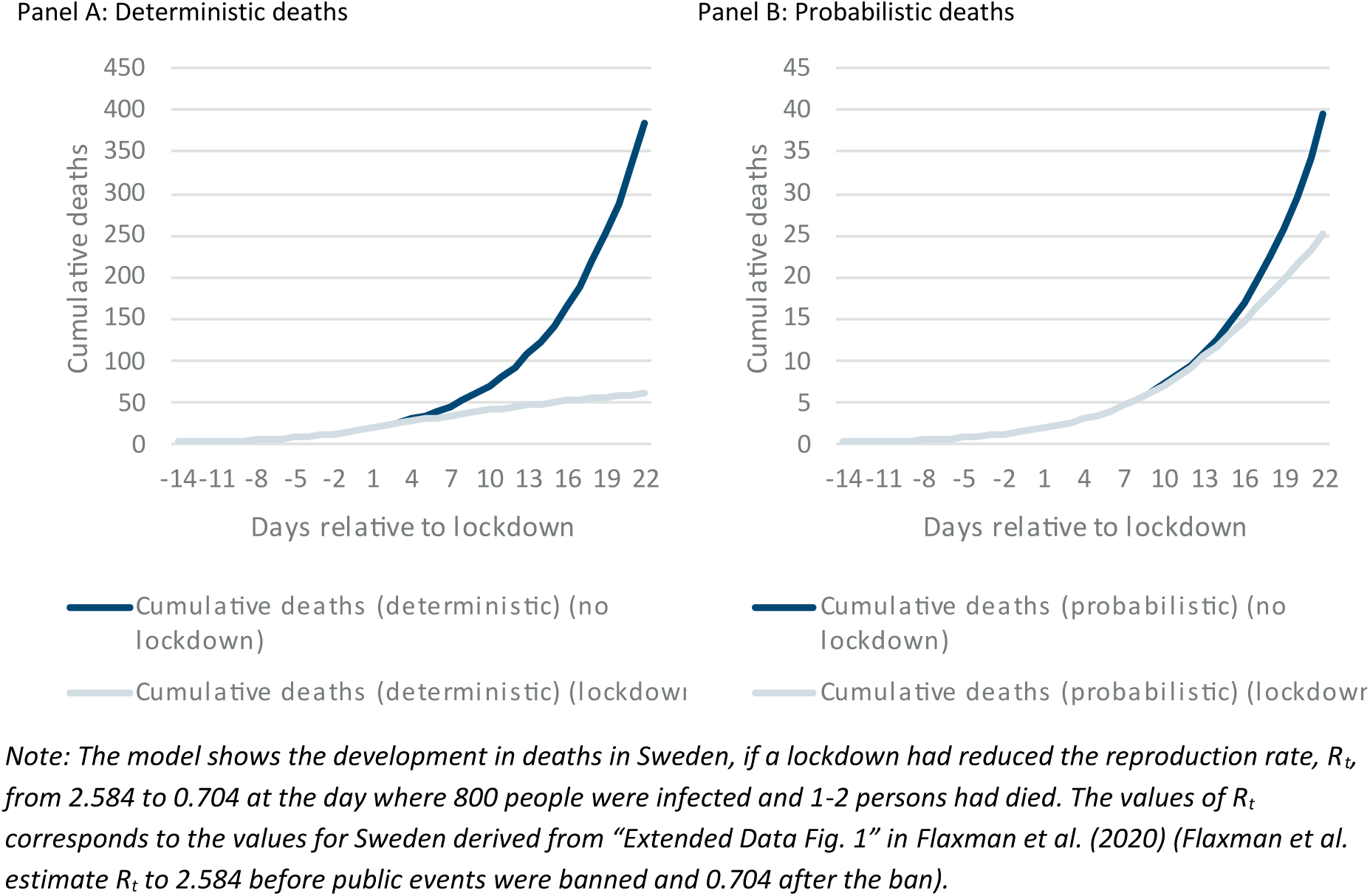
Fatal disease duration distribution from infection to death.

As Wood (2022) illustrate^18^, R_t_ changed gradually as governments informed the public and advised people so self-isolate and work from home. Figure 15 below illustrates the importance of this insight for when the effect of a lockdown is visible in cumulative COVID-19 deaths. The figure shows the relative difference (deaths with lockdown relative to deaths without lockdown) for different values of R_t_ before lockdown brings R_t_ down to 0.704. The figure shows that if R_t_ = 1.5 before lockdown, the difference in cumulative deaths with and without an effective lockdown is only approximately 8% after 21 days. If R_t_ = 2.0 the difference after 21 days is 15%, and if R_t_ = 2.584 the difference is 32%.

**Figure 15.**
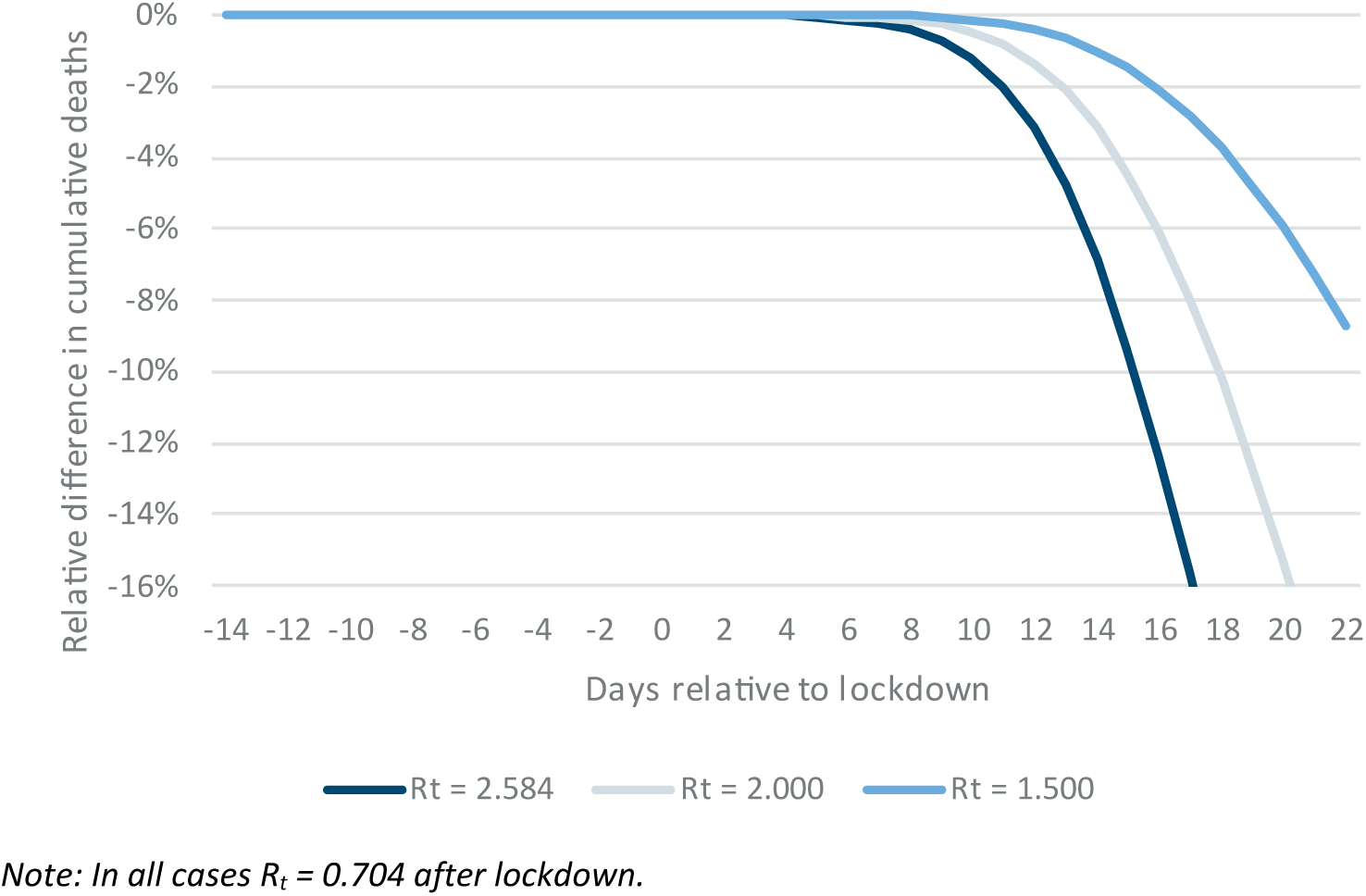
Difference in cumulative deaths given different values of Rt prior to lockdown.

In all cases there is virtually no difference after 12 days, and after 15 days the difference is less than 10%.

## Supplementary material

R-code available at https://github.com/JonasHerby/DontJumptoFaultyConclusions

## Footnotes

1 See Appendix IV in Herby et al. (2023).

2 For example, Born et al. (2021) estimate that “a lockdown would have reduced the number of deaths by 38%, from 5,795 to 3,573” (2,222 avoided deaths), Ege et al. (2023) estimate that “up to about 4411” deaths could have been avoided, and Conyon and Thomsen (2021)’s estimate corresponds to app. 3,000 avoided deaths. These numbers translate to approximately 1,250 to 2,500 avoided deaths in Denmark with a population almost half of that in Sweden. The difference-in-difference studies estimate similar effects of lockdowns. In comparison, influenza causes an excess mortality of 1,644 deaths in the 2017/18 influenza season according to Statens Serum Institut (2018).

3 Week 6: February 3-9, Week 7: February 10-16, Week 8: February 17-23, Week 9: March 2-8, Week 10: March 9-15.

4 See https://www.mortality.org/File/GetDocument/Public/STMF/DOC/STMFNote.pdf p. 6.

5 Indicator id’s SH.MED.BEDS.ZS, SP.URB.TOTL.IN.ZS, NY.GDP.PCAP.PP.KD, SP.POP.65UP.TO.ZS, and SM.POP.TOTL.ZS.

6 We use the tables “Döda efter region, vecka och ålder” and “Folkmängd efter region, år och ålder”. Links to all datasets are available in our R-code provided as supplementary material.

7 Taiwan is not part of the donor pool as the World Bank does not supply data for Taiwan. Also, Russia is not part of the donor pool as there are no mortality data after week 53 of 2020.

8 See supplementary information for Arnarson (2021), Table C1

9 AUS, AUT, BEL, BGR, CAN, CHE, CHL, CZE, DEU, DNK, ESP, EST, FIN, FRA, GRC, HRV, HUN, ISL, ISR, ITA, KOR, LTU, LUX, LVA, NLD, NOR, NZL, POL, PRT, SVK, SVN, SWE, USA, and NIR, SCO & ENW from GBR

10 See Figure 2, panel a, in Ege et al. (2023).

11 Choosing 2011 excludes Australia, Chile, Northern Ireland, Greece and USA from the donor pool. Choosing 2007 further excludes Italy, New Zealand, Great Britain (England-Wales), and Korea.

12 See for example Bussolo et al. (2023).

13 The donors are Netherland (37%), Canada (30%), Finland (23%), Denmark (9%), and Luxembourg (1%). The distribution of vaccines in Sweden was comparable to or slower than the distribution of vaccines in the donor countries. See https://ourworldindata.org/explorers/coronavirus-data-explorer?zoomToSelection=true&time=earliest..2021-06-30&facet=none&pickerSort=asc&pickerMetric=location&hideControls=true&Metric=People+vaccinated&Interval=Cumulative&Relative+to+Population=true&Color+by+test+positivity=false&country=SWE∼CAN∼FIN∼NLD∼DNK and https://ourworldindata.org/explorers/coronavirus-data-explorer?zoomToSelection=true&time=earliest..2021-06-30&facet=none&country=SWE∼CAN∼FIN∼NLD∼DNK&pickerSort=asc&pickerMetric=location&hideControls=true&Metric=Vaccine+doses&Interval=Cumulative&Relative+to+Population=true&Color+by+test+positivity=false

14 Sebhatu et al. (2020) finds a statistically significant correlation between COVID-19 mortality and the degree of lockdown in a country, but the magnitude is modest, explaining only 2.1 stringency points on average. In comparison, the gap between the strictest and most lenient lockdowns in Europe was between 67 and 92 stringency points in the period from 16 March to 15 April 2020), Herby et al. (2023), 117.

15 See Appendix IV in Herby et al. (2023).

16 García-García et al. (2021) find a somewhat different distribution, where app. 20% dies within 14 days (Wood (2022): 11%), 42% dies on day 15 to 21 (29%), and 25% on day 22 to 28 (26%). However, the overall conclusion, that the effect of lockdowns are not visible in mortality data the first 14 days, remains. e

17 The SEIR-model was downloaded from https://www.wku.edu/da/covid-19-research/excel_sir_model.php (direct link: https://www.wku.edu/da/covid-19-research/sir_model.xlsx) and used the following values: Gamme = 0.2, Delta = 0.005, Lambda = 0.333 and Beta = 0,5168 (before lockdown) and 0,1408 (after lockdown). The values of Beta corresponds to the values of the reproduction rate, Rt, for Sweden derived from “Extended Data Fig. 1” in Flaxman et al. (2020) (they estimate Rt to 2.584 before public events were banned and 0.704 after the ban).

18 For example, see Wood (2022), Figure 3.

## References

Abadie, Alberto. 2021. “Using Synthetic Controls: Feasibility, Data Requirements, and Methodological Aspects.” Journal of Economic Literature 59 (2):391–425. 10.1257/jel.20191450.

Abadie, Alberto, Alexis Diamond, and Jens Hainmueller. 2010. “Synthetic Control Methods for Comparative Case Studies: Estimating the Effect of California’s Tobacco Control Program.” Journal of the American Statistical Association 105 (490). Taylor & Francis:493–505. 10.1198/jasa.2009.ap08746.

Abadie, Alberto, Alexis Diamond, and Jens Hainmueller. 2015. “Comparative Politics and the Synthetic Control Method.” American Journal of Political Science 59 (2). John Wiley & Sons, Ltd:495–510. 10.1111/ajps.12116.

Abadie, Alberto, and Javier Gardeazabal. 2003. “The Economic Costs of Conflict: A Case Study of the Basque Country.” American Economic Review 93 (1):113–32. 10.1257/000282803321455188.

Arnarson, Björn Thor. 2021. “How a School Holiday Led to Persistent COVID-19 Outbreaks in Europe.” Scientific Reports 11 (1):24390. 10.1038/s41598-021-03927-z.

Björk, Jonas, Kristoffer Mattisson, and Anders Ahlbom. 2021. “Impact of Winter Holiday and Government Responses on Mortality in Europe during the First Wave of the COVID-19 Pandemic.” European Journal of Public Health 31 (2):272–77. 10.1093/eurpub/ckab017.

Bjørnskov, Christian, and Stefan Voigt. 2022. “This Time Is Different?—On the Use of Emergency Measures during the Corona Pandemic.” European Journal of Law and Economics 54 (1):63–81. 10.1007/s10657-021-09706-5.

Born, Benjamin, Alexander M. Dietrich, and Gernot J. Müller. 2021. “The Lockdown Effect: A Counterfactual for Sweden.” PLOS ONE 16 (4). Public Library of Science:e0249732. 10.1371/journal.pone.0249732.

Bussolo, Maurizio, Nayantara Sarma, and Iván Torre. 2023. “The Links between COVID-19 Vaccine Acceptance and Non-Pharmaceutical Interventions.” Social Science & Medicine 320 (March):115682. 10.1016/j.socscimed.2023.115682.

Cho, Sang-Wook (Stanley). 2020. “Quantifying the Impact of Nonpharmaceutical Interventions during the COVID-19 Outbreak: The Case of Sweden.” The Econometrics Journal 23 (3):323–44. 10.1093/ectj/utaa025.

Conyon, Martin J., Lerong He, and Steen Thomsen. 2020. “Lockdowns and COVID-19 Deaths in Scandinavia.” *SSRN Electronic Journal*, June. 10.2139/ssrn.3616969.

Conyon, Martin J., and Steen Thomsen. 2021. “COVID-19 in Scandinavia.” 10.2139/ssrn.3793888.

Ege, Florian, Giovanni Mellace, and Seetha Menon. 2023. “The Unseen Toll: Excess Mortality during Covid-19 Lockdowns.” Scientific Reports 13 (1):18745. 10.1038/s41598-023-45934-2.

Engler, Sarah, Palmo Brunner, Romane Loviat, Tarik Abou-Chadi, Lucas Leemann, Andreas Glaser, and Daniel Kübler. 2021. “Democracy in Times of the Pandemic: Explaining the Variation of COVID-19 Policies across European Democracies.” West European Politics 44 (5–6). Routledge:1077–1102. 10.1080/01402382.2021.1900669.

Ferguson, Neil M, Daniel Laydon, Gemma Nedjati-Gilani, Natsuko Imai, Kylie Ainslie, Marc Baguelin, Sangeeta Bhatia, et al. 2020. “Impact of Non-Pharmaceutical Interventions (NPIs) to Reduce COVID-19 Mortality and Healthcare Demand,” March, 20.

Flaxman, Seth, Swapnil Mishra, Axel Gandy, H. Juliette T. Unwin, Thomas A. Mellan, Helen Coupland, Charles Whittaker, et al. 2020. “Estimating the Effects of Non-Pharmaceutical Interventions on COVID-19 in Europe.” Nature 584 (7820):257–61. 10.1038/s41586-020-2405-7.

García-García, David, María Isabel Vigo, Eva S. Fonfría, Zaida Herrador, Miriam Navarro, and Cesar Bordehore. 2021. “Retrospective Methodology to Estimate Daily Infections from Deaths (REMEDID) in COVID-19: The Spain Case Study.” Scientific Reports 11 (1). Nature Publishing Group:11274. 10.1038/s41598-021-90051-7.

Gordon, Daniel V., R. Quentin Grafton, and Stein Ivar Steinshamn. 2020. “Statistical Analyses of the Public Health and Economic Performance of Nordic Countries in Response to the COVID-19 Pandemic,” November, 2020.11.23.2023 711. 10.1101/2020.11.23.20236711.

Grier, Kevin, and Norman Maynard. 2016. “The Economic Consequences of Hugo Chavez: A Synthetic Control Analysis.” Journal of Economic Behavior & Organization 125 (May):1–21. 10.1016/j.jebo.2015.12.011.

Herby, Jonas, Lars Jonung, and Steve H. Hanke. 2023. Did Lockdowns Work? The Verdict on Covid Restrictions. IEA Perspectives 1/IEA. London: IEA. https://iea.org.uk/wp-content/uploads/2023/06/Perspectives-_1_Did-lockdowns-work June_web-1.pdf.

Juranek, Steffen, and Floris T. Zoutman. 2021. “The Effect of Non-Pharmaceutical Interventions on the Demand for Health Care and on Mortality: Evidence from COVID-19 in Scandinavia.” Journal of Population Economics 34 (4):1299–1320. 10.1007/s00148-021-00868-9.

Latour, Chiara, Franco Peracchi, and Giancarlo Spagnolo. 2022. “Assessing Alternative Indicators for Covid-19 Policy Evaluation, with a Counterfactual for Sweden.” PLOS ONE 17 (3). Public Library of Science:e0264769. 10.1371/journal.pone.0264769.

Mader, Sebastian, and Tobias Rüttenauer. 2021. “The Effects of Non-Pharmaceutical Interventions on COVID-19-Related Mortality: A Generalized Synthetic Control Approach across 169 Countries.” SocArXiv. 10.31235/osf.io/v2ef8.

Mader, Sebastian, and Tobias Rüttenauer. 2022. “The Effects of Non-Pharmaceutical Interventions on COVID-19 Mortality: A Generalized Synthetic Control Approach Across 1 9 Countries.” Frontiers in Public Health 10 (April):820642. 10.3389/fpubh.2022.820642.

Mistur, Evan M., John Wagner Givens, and Daniel C. Matisoff. 2023. “Contagious COVID 19 Policies: Policy Diffusion during Times of Crisis.” Review of Policy Research 40 (1):36–62. 10.1111/ropr.12487.

Mulligan, Casey B., and Robert D. Arnott. 2022. “The Young Were Not Spared: What Death Certificates Reveal about Non-Covid Excess Deaths.” INQUIRY: The Journal of Health Care Organization, Provision, and Financing 59 (January). SAGE Publications Inc:00469580221139016. 10.1177/00469580221139016.

Our World in Data. 2022. “COVID-19 Data Explorer.” Our World in Data. 2022. https://ourworldindata.org/coronavirus-data-explorer.

Sebhatu, Abiel, Karl Wennberg, Stefan Arora-Jonsson, and Staffan I. Lindberg. 2020. “Explaining the Homogeneous Diffusion of COVID-19 Nonpharmaceutical Interventions across Heterogeneous Countries.” PNAS, August, 202010625. 10.1073/pnas.2010625117.

Statens Serum Institut. 2018. “EPI_NYT Uge 23/24 - 2018.” June 13, 2018. https://www.ssi.dk/aktuelt/nyhedsbreve/epi-nyt/2018/uge-23_24---2018.

Tegnell, Anders. 2023. Tankar efter en pandemi: Och lärdomarna inför nästa. Natur & Kultur Digital.

Wood, Simon N. 2022. “Inferring UK COVID-19 Fatal Infection Trajectories from Daily Mortality Data: Were Infections Already in Decline before the UK Lockdowns?” Biometrics 78 (3):1127–40. 10.1111/biom.13462.

