## Supplementary material for "Don’t Jump to Faulty Conclusions: Using the Synthetic Control Method to Evaluate the Effect of a Counterfactual Lockdown in Sweden": R-code: 010 Create Mortality_data.R

```
# library(Synth)
# library(gapminder)
# library(tidyverse)
# library(cowplot)
library(ggplot2)
# library(ggplotify)
# library(grid)
# library(ggpubr)
# library(gridGraphics)
# library(scpi)
# library(augsynth)
# library(synthdid)
library(readxl)
# library(xtable)
library(dplyr)
library(tidyr)
# library(ISOweek)
# library(purrr)
library(openxlsx)
# library(scales)
library(zoo)

rm(list=ls())
setwd(dirname(rstudioapi::getActiveDocumentContext()$path))
cat("\014")  #Clearer console


# The final data have been created from the following data sources:
# Mortality data (Short-Term Mortality Fluctuations) https://www.mortality.org/Data/STMF
# GDP per capita 2020 (Worldbank) https://ourworldindata.org/grapher/gdp-per-capita-worldbank?tab=chart&country=AUT
# Age-standardized death rate from cardiovascular diseases 2019 (Our World in Data) https://ourworldindata.org/grapher/age-standardized-death-rate-cardiovascular-disease?tab=chart&country=AUT
# Life expectancy 2019 (Worldbank) https://ourworldindata.org/grapher/life-expectancy-at-birth-total-years?tab=chart&country=AUT
# Share of people living in urban agglomerations of more than 1 million 2020 (Our World in Data) https://ourworldindata.org/grapher/share-of-population-urban?tab=chart&time=latest&country=AUT
# Population density 2020 (Our World in Data) https://ourworldindata.org/grapher/population-density?tab=chart&time=1851..2022&country=~AUT
# Links accessed on 10-10-2023


# Purpose -----------------------------------------------------------------

# We want to check the robustness of the results in "The unseen toll: excess mortality during covid-19 lockdowns" (Ege et al. 2023) by
# 1) Fitting the SCM on a much longer data period prior to lockdowns. (Ege et al. 2023) fit their model based on the period starting 
#    first week of November 2019. We would like to fit it based on several years.
# 2) Calculate expected mortality ignoring the 2018/19 winter which was especially easy on Swedish elderly (very few deaths)
# 3) Limiting the donor pool to countries on the Northern Hemisphere (similar to excluding New Zealand from the donor pool), 
#    secondly European countries (excluding Israel, South Korea, New Zealand and Canada).
# 4) Handle the impact of Winter holiday in week 9 which is known to have had a large impact on COVID-19


# Prepare datasets -------------------------------------------------------------

## Load mortality data from mortality.org data and create new variables ---------------------------------

# url <- "https://www.mortality.org/File/GetDocument/Public/STMF/Outputs/stmf.csv"
# destfile <- "stmf.csv"  # Specify your desired path here
# download.file(url, destfile, method="curl")  # 'method="curl"' might be necessary on some platforms

stmf <- read.csv("stmf.csv", skip = 2)
stmf_data <- stmf[stmf$Sex == "b",]  #Only keep "both" sexes

# Check if there is a full dataset with data for all age groups
# Calculate the number of non-NA values for each specified variable by country
non_na_counts <- stmf_data %>%
  group_by(CountryCode) %>%
  summarise(
    NonNA_DTotal = sum(!is.na(DTotal)),
    NonNA_D0_14 = sum(!is.na(D0_14)),
    NonNA_D15_64 = sum(!is.na(D15_64)),
    NonNA_D65_74 = sum(!is.na(D65_74)),
    NonNA_D75_84 = sum(!is.na(D75_84)),
    NonNA_D85p = sum(!is.na(D85p))
  ) %>%
  # Add a logical check to see if all non-NA counts are the same for each country
  mutate(SameNonNA = NonNA_DTotal == NonNA_D0_14 & NonNA_D0_14 == NonNA_D15_64 & 
           NonNA_D15_64 == NonNA_D65_74 & NonNA_D65_74 == NonNA_D75_84 & NonNA_D75_84 == NonNA_D85p)

# Check if there's any FALSE in SameNonNA column
if(any(!non_na_counts$SameNonNA)) {
  stop("Error: The numbers of non-NAs are not the same in each row.")
} else {
  cat("QA OK! All rows have the same number of non-NAs across variables.")
}

### Calculate populations sizes ---------------------------------------------
# See https://www.mortality.org/File/GetDocument/Public/STMF/DOC/STMFNote.pdf -- 
# the part after "The weekly age-specific death rates are calculated as follows:"
stmf_data$PopulationTotal <- stmf_data$DTotal / stmf_data$RTotal * 52

# For some country/weeks the rate is zero, so we cannot calculate population size based on weekly data.
# Instead we find a representative obs for each country/year where all rates are >0 and calculate the 
# country/year population on this basis.

# Step 1 & 2: Filter for rows without NAs in rate columns and select a representative row
population_estimates <- stmf_data %>%
  filter(R0_14>0 & R15_64>0 & R65_74>0 & R75_84>0 & R85p>0) %>%
  group_by(CountryCode, Year) %>%
  slice(1) %>%  # Assuming the first row is representative
  ungroup() %>%
  # Step 3: Calculate population sizes
  mutate(
    Population0_14 = D0_14 / R0_14 * 52,
    Population14_64 = D15_64 / R15_64 * 52,
    Population65_74 = D65_74 / R65_74 * 52,
    Population75_84 = D75_84 / R75_84 * 52,
    Population85p = D85p / R85p * 52
  ) %>%
  # Select only necessary columns to merge
  select(CountryCode, Year, Population0_14, Population14_64, Population65_74, Population75_84, Population85p)

# Step 4: Merge the population estimates back onto the original dataframe
stmf_data <- left_join(stmf_data, population_estimates, by = c("CountryCode", "Year"))

#Population QA
stmf_data <- stmf_data %>% 
  mutate(PopulationQA = PopulationTotal - Population0_14 - Population14_64 - Population65_74 - Population75_84 - Population85p)

# Check if there's any large differences
if(abs(max(stmf_data$PopulationQA)) + abs(min(stmf_data$PopulationQA)) > 1) {
  stop("Error: The sum of age groups populations does not equal total population")
} else {
  cat("QA OK! Populations are calculated correctly")
}
stmf_data$PopulationQA <- NULL

### Generate new age groups ---------------------------------------------
#Population
stmf_data <- stmf_data %>%
  mutate(Population0_64 = Population0_14 + Population14_64,
         Population65p = Population65_74 + Population75_84 + Population85p)

#Deaths
stmf_data <- stmf_data %>%
  mutate(D0_64 = D0_14 + D15_64, 
         D65p = D65_74 + D75_84 + D85p) 

#Death rate
stmf_data <- stmf_data %>%
  mutate(R0_64 = D0_64 / Population0_64 * 52,
         R65p = D65p / Population65p * 52)


#Weekly death rate for DTotal
stmf_data$RTotal_weekly <- stmf_data$RTotal / 52
stmf_data$R0_64_weekly <- stmf_data$R0_64 / 52
stmf_data$R65p_weekly <- stmf_data$R65p / 52

### Clean data and remove unused data frames --------------------------------
#rm(non_na_counts, population_estimates, stmf)

## Load Swedish mortality and holiday data split on län ------------------------

### Load mortality data --------------------------------------------------------
# url <- "https://www.statistikdatabasen.scb.se/sq/151069"
# destfile <- "SWE_mortality.xlsx"  # Specify your desired path here
# download.file(url, destfile, method="curl")  # 'method="curl"' might be necessary on some platforms

SWE_mortality <- read_excel("SWE_mortality.xlsx", skip = 2)
SWE_mortality <- SWE_mortality %>% mutate(`...1` = zoo::na.locf(`...1`))
# mutate(...1 = zoo::na.locf(...1)): The mutate function modifies the SWE_mortality 
# data frame. Inside mutate, I'm using the zoo::na.locf() function from the zoo package
# to fill in NA values, as län names are only listed for the first obs.
# na.locf() stands for "Last Observation Carried Forward" and 
# replaces each NA with the most recent non-NA value. If the first value is NA, it
# remains NA because there's no previous value to carry forward.
colnames(SWE_mortality)[1] <- "Region"
colnames(SWE_mortality)[2] <- "YearWeek"
SWE_mortality$YearWeek <- gsub("V", "", SWE_mortality$YearWeek)
SWE_mortality$Year <- as.numeric(substr(SWE_mortality$YearWeek, 1, 4))
SWE_mortality$Week <- as.numeric(substr(SWE_mortality$YearWeek, 5, 6))
SWE_mortality$YearWeek <- as.numeric(SWE_mortality$YearWeek)
#Drop data from 2024
SWE_mortality <- SWE_mortality %>%
  filter(Year < 2024)

SWE_mortality <- SWE_mortality %>%
  mutate(
    DTotal = totalt,
    D0_60 = `0-34 år` + `35-59 år`,
    D60_69 = `60-69 år`,
    D70p = `70-79 år` + `80-89 år` + `90+ år`
  )

#### Synthesize D0_65 & D65p

SWE_synth <- SWE_mortality %>%
  group_by(Year, Week) %>%
  summarise(
    DTotal = sum(DTotal, na.rm = TRUE),
    D0_60 = sum(D0_60, na.rm = TRUE),
    D60_69 = sum(D60_69, na.rm = TRUE),
    D70p = sum(D70p, na.rm = TRUE),
    .groups = "drop"  # Ensures ungrouping after summarise
  )

stmf_synth <- stmf_data %>%
  filter(CountryCode == "SWE") %>%
  select(Year, Week, DTotal, D0_64, D65p) %>%
  rename(DTotal_stmf = DTotal)

synth_data <- left_join(SWE_synth, stmf_synth, by=join_by(Year, Week))

synth_data <- synth_data %>%
  mutate(
    D60_64 = D0_64 - D0_60,
    share_60_64_of_60_69 = D60_64 / D60_69
  )

SWE_mortality <- left_join(SWE_mortality, synth_data %>% select(Year, Week, share_60_64_of_60_69), by=join_by(Year, Week))

SWE_mortality <- SWE_mortality %>%
  mutate(
    D0_64 = D0_60 + D60_69 * share_60_64_of_60_69,
    D65p = D70p + D60_69 * (1-share_60_64_of_60_69)
  )


### Load population data -------------------------------------------------------
# url <- "https://www.statistikdatabasen.scb.se/sq/151070"
# destfile <- "SWE_population.xlsx"  # Specify your desired path here
# download.file(url, destfile, method="curl")  # 'method="curl"' might be necessary on some platforms

SWE_population <- read_excel("SWE_population.xlsx", skip = 2)
#Remove notes etc. in bottom of file by filtering out obs where column 2 is NA
SWE_population <- SWE_population %>%
  filter(!is.na(.[[2]]))
SWE_population <- SWE_population %>%
  mutate(`...1` = zoo::na.locf(`...1`))
colnames(SWE_population)[1] <- "Region"
colnames(SWE_population)[2] <- "Year"
SWE_population$Year <- as.numeric(SWE_population$Year)

#Create same age groups as for mortality
SWE_population <- SWE_population %>%
  mutate(
    Population0_64 = `0-4 år` + `5-9 år` + `10-14 år` + `15-19 år` + 
      `20-24 år` + `25-29 år` + `30-34 år` + `35-39 år` + `40-44 år` + 
      `45-49 år` + `50-54 år` + `55-59 år` + `60-64 år`,
    Population65p = `65-69 år` + `70-74 år` + `75-79 år` + `80-84 år` + 
      `85-89 år` + `90-94 år` + `95-99 år` + `100+ år`,
    PopulationTotal = Population0_64 + Population65p
  )

### Create 'regions' based on winter holiday week ---------------------

# Creating the data frame with the region number and name, holiday week, and whether it is a former Danish region
# Source for holiday weeks: https://www.nature.com/articles/s41598-021-03927-z
Region_data <- data.frame(
  Region = c("01 Stockholms län", "03 Uppsala län", "04 Södermanlands län",
             "05 Östergötlands län", "06 Jönköpings län", "07 Kronobergs län",
             "08 Kalmar län", "09 Gotlands län", "10 Blekinge län",
             "12 Skåne län", "13 Hallands län", "14 Västra Götalands län",
             "17 Värmlands län", "18 Örebro län", "19 Västmanlands län",
             "20 Dalarnas län", "21 Gävleborgs län", "22 Västernorrlands län",
             "23 Jämtlands län", "24 Västerbottens län", "25 Norrbottens län"),
  HolidayWeek_org = c("w9", "w8", "w8", "w8", "w7", "w8", "w8", "w9", "w8", "w8", "w8", "w7", "w9", "w8", "w9", "w9", "w9", "w10", "w10", "w10", "w10"), # Original
  HolidayWeek_com =  c("w910", "w78", "w78", "w78", "w78", "w78", "w78", "w910", "w78", "w78", "w78", "w78", "w910", "w78", "w910", "w910", "w910", "w910", "w910", "w910", "w910"), # 7+78 combined to 78 & 910+10 combined to 910
  #  HolidayWeek_adj =  c("adjw910", "adjw910", "adjw910", "adjw78", "adjw78", "adjw78", "adjw78", "adjw910", "adjw78", "adjw78", "adjw78", "adjw78", "adjw910", "adjw78", "adjw910", "adjw910", "adjw910", "adjw910", "adjw910", "adjw910", "adjw910"), # 7+78 combined to 78 & 910+10 combined to 910", but "03 Uppsala län" & "04 Södermanlands län" put in the week 910 category
  #  FormerDanish = c("fSE", "fSE", "fSE", "fSE", "fSE", "fSE", "fSE", "fSE", "fDK", "fDK", "fDK", "fSE", "fSE", "fSE", "fSE", "fSE", "fSE", "fSE", "fSE", "fSE", "fSE"),  #DK = Former Danish Region
  Sweden_SCB = c("SCB", "SCB", "SCB", "SCB", "SCB", "SCB", "SCB", "SCB", "SCB", "SCB", "SCB", "SCB", "SCB", "SCB", "SCB", "SCB", "SCB", "SCB", "SCB", "SCB", "SCB") # This is used to compare to Mortality.org data later
)


### Create Swedish mortality data set corresponding to stmf_data ----------------
#Subset data and rename columns
# Assuming you want to keep only some columns and rename 'totalt' to 'DTotal'
SWE_mort_data <- SWE_mortality %>%
  select(
    Region,
    Year,
    Week,
    YearWeek,
    DTotal,
    D0_64,
    D65p
  )

SWE_pop_data <- SWE_population %>%
  select(
    Region,
    Year,
    PopulationTotal,
    Population0_64,
    Population65p
  )

SWE_data <- left_join(SWE_mort_data, SWE_pop_data, by=c("Region", "Year"))
SWE_data <- left_join(SWE_data, Region_data, by="Region")

#### Calculate share 60+ and 70+ in each region group ------
# Group by Year and HolidayWeek_org, then summarize the ratio
summary_data <- SWE_data %>%
  group_by(Year, HolidayWeek_org) %>%
  summarize(Ratio65p = sum(Population65p, na.rm = TRUE) / sum(PopulationTotal, na.rm = TRUE),
            .groups = "drop"
  )

# Pivot the data to wide format
wide_data65p <- summary_data %>%
  pivot_wider(names_from = HolidayWeek_org, values_from = Ratio65p) %>%
  select(Year, w7, w8, w9, w10)

# Display the wide data
print(wide_data65p)
write.xlsx(wide_data65p, file = "Share of population 65+.xlsx")

### Create mortality data grouped on holiday week ---------------------------
create_swedens <- function(df, column) {
  df %>%
    group_by(!!sym(column), Year, Week) %>%
    summarize(
      DTotal = sum(DTotal, na.rm = TRUE),
      D0_64 = sum(D0_64, na.rm = TRUE),
      D65p = sum(D65p, na.rm = TRUE),
      PopulationTotal = sum(PopulationTotal, na.rm = TRUE),
      Population0_64 = sum(Population0_64, na.rm = TRUE),
      Population65p = sum(Population65p, na.rm = TRUE),
      .groups = "drop"
    ) %>%
    mutate(Region = paste0("SWE_", !!sym(column)),
           ColName = column) %>%
    select(-!!sym(column))
}

# List of columns to process
region_cols <- names(Region_data[,2:length(Region_data)])

# Apply the function to each holiday week column and combine the results
SWE_data_groups <- bind_rows(
  lapply(region_cols, function(col) create_swedens(SWE_data, col))
)


# QA: Summarize DTotal by Region
sum_DTotal <- sum(SWE_data$DTotal)
sum_PopulationTotal <- sum(SWE_data$PopulationTotal)

SWE_data_summary <- SWE_data_groups %>%
  group_by(ColName) %>%
  summarize(
    Total_DTotal = sum(DTotal, na.rm = TRUE),
    Total_Population = sum(PopulationTotal, na.rm = TRUE),
    Diff_DTotal = Total_DTotal - sum_DTotal,
    Diff_PopTotal = Total_Population - sum_PopulationTotal,
    .groups = "drop"
  )

if(any(SWE_data_summary$Diff_DTotal != 0) | any(SWE_data_summary$Diff_PopTotal != 0)){stop("Check data!")}

SWE_data_groups$ColName <- NULL

# Calculate mortality rate
SWE_data_groups <- SWE_data_groups %>%
  mutate(
    RTotal_weekly = DTotal / PopulationTotal,
    R0_64_weekly = D0_64 / Population0_64,
    R65p_weekly = D65p / Population65p
  )

#SWE_data_groups$RTotal <- SWE_data_groups$RTotal_weekly * 52
# See https://www.mortality.org/File/GetDocument/Public/STMF/DOC/STMFNote.pdf -- 
# the part after "The weekly age-specific death rates are calculated as follows:"

# Rename Regions to CountryCode so it corresponds to mortality.org data
SWE_data_groups <- SWE_data_groups %>%
  rename(CountryCode = Region)

### Clean data and remove unused df's---------------
# rm(SWE_data, Region_data, SWE_data_summary)
# rm(SWE_mort_data, SWE_mortality, SWE_pop_data, SWE_population)


# Combine stmf_data and SWE_data_groups ---------------------------------------

# YearWeekMin <- 201601 # The cut-off date for counrties in the data set. All countries which do not have data from this YearWeek and forward are excluded
# MyYear <- floor(YearWeekMin/100)
# MyWeek <- YearWeekMin-MyYear*100

#Combine data but only keep columns (i.e. age groups) available in SWE_data_groups
Mortality_data <- bind_rows(stmf_data %>% select(c(names(SWE_data_groups))), SWE_data_groups)


##Generate additional variables
#Generate YearWeek-variable
Mortality_data$YearWeek <- Mortality_data$Year*100+Mortality_data$Week

#Generate weekly mortality rates per 100,000
Mortality_data$RTotal_weekly100K <- Mortality_data$RTotal_weekly*100000
Mortality_data$R0_64_weekly100K <- Mortality_data$R0_64_weekly *100000
Mortality_data$R65p_weekly100K <- Mortality_data$R65p_weekly *100000


# Reorganize columns ------------------------------------------------------
Mortality_data <- Mortality_data %>% 
  mutate(CountryCode = ifelse(CountryCode == "SWE", "SWE_STMF", CountryCode)) %>%
  mutate(CountryCode = ifelse(CountryCode == "SWE_SCB", "SWE", CountryCode))

# Summarize populations ---------------------------------------------------


### Summarize PopulationTotal by CountryCode for CountryCodes starting with "SWE" and YearWeek == 202001 ----------
summary_population <- Mortality_data %>%
  filter(grepl("^SWE", CountryCode) & YearWeek == 202001) %>%
  group_by(CountryCode) %>%
  summarize(TotalPopulation = sum(PopulationTotal, na.rm = TRUE))

# Print and save the summary
print(summary_population)
write.xlsx(summary_population, file = paste0("Population summary SWE.xlsx"))


Mortality_data <- Mortality_data %>% 
  select(
    CountryCode,
    Year,
    Week,
    YearWeek,
    RTotal_weekly100K,
    R0_64_weekly100K,
    R65p_weekly100K
  ) %>% filter(YearWeek >= 200427 & YearWeek <= 202326)

# Save data ---------------------------------------------------------------

save(Mortality_data, file = "Mortality_data.RData")
```
