## Supplementary material for "Don’t Jump to Faulty Conclusions: Using the Synthetic Control Method to Evaluate the Effect of a Counterfactual Lockdown in Sweden": R-code: 020 Create WB_data.R

```
# Load the WDI package
library(WDI)
library(purrr)
library(dplyr)

search_indicators <- WDIsearch(string = "65 and above")

indicators <- list()
indicators[[1]] <- c("SH.MED.BEDS.ZS",     "Hospital_beds",    "Hospital beds (per 1,000 people)")
indicators[[2]] <- c("SP.URB.TOTL.IN.ZS",  "Urban population", "Urban population (% of total population)")
indicators[[3]] <- c("NY.GDP.PCAP.PP.KD",  "GDPCAP_PPP",       "GDP per capita, PPP (constant 2021 international $)")
indicators[[4]] <- c("SP.POP.65UP.TO.ZS",  "ShareAged65p",     "Population ages 65 and above (% of total population)")
indicators[[5]] <- c("SM.POP.TOTL.ZS",     "Migrant_share",    "International migrant stock (% of population)")

#Manually test if all data sets are complete
# data <- WDI(indicator = indicators[[1]][1], start = 2000, end = 2023, extra = TRUE)
# nrow(data)


# Function to fetch data from WB and prepare it for merging
fetch_and_prepare_data <- function(indicator) {
  data <- WDI(indicator = indicator[1], start = 2000, end = 2023, extra = TRUE) %>%
    select(iso3c, year, !!sym(indicator[1])) %>%
    filter(!is.na(iso3c) & iso3c !="") %>%
    rename(CountryCode = iso3c,
           Year = year)
  colnames(data)[3] <- indicator[2] # Rename the third column to the short name
  return(data)
}


# Fetch data for all indicators and store in a list
data_list <- map(indicators, fetch_and_prepare_data)

# Merge all data frames in the list into one data frame
WB_data <- reduce(data_list, left_join, by = c("CountryCode", "Year"))

save(WB_data, file = "WB_data.RData")
```
