## Supplementary material for "Don’t Jump to Faulty Conclusions: Using the Synthetic Control Method to Evaluate the Effect of a Counterfactual Lockdown in Sweden": R-code: 100 SCM, lockdown and winter holidays.r

```
library(Synth)
# library(gapminder)
# library(tidyverse)
# library(cowplot)
library(ggplot2)
# library(ggplotify)
# library(grid)
# library(ggpubr)
# library(gridGraphics)
# library(scpi)
# library(augsynth)
# library(synthdid)
library(readxl)
# library(xtable)
library(dplyr)
library(tidyr)
# library(ISOweek)
# library(purrr)
library(openxlsx)
# library(scales)
library(stringr)
library(zoo)

rm(list=ls())
setwd(dirname(rstudioapi::getActiveDocumentContext()$path))
cat("\014")  #Clearer console


# Purpose -----------------------------------------------------------------

#Set this to TRUE to skip the code and only update plots
onlyPlots <- FALSE

# Load and prepare data -------------------------------------------------------------

## Define variables etc. which will create new folders to store results -----

#Set periods to analyze
prewindows <- c(201627, 201127, 200727)

# Choose the end of the analyzed period
postwindow <- 202126

#set list of predictors to analyze
mypredictors <- c("full", "single")
mypredictors <- c("full") #We only use the full predictor set

#set list of dependents to analyze
dependents <- c("RTotal_weekly100K", "R0_64_weekly100K", "R65p_weekly100K")
dependents <- c("RTotal_weekly100K") #We only look at the total mortality

#Set list of SWE-related country codes to analyze
swe_related_codes <- c("SWE", "SWE_w78", "SWE_w910", "SWE_w7", "SWE_w8", "SWE_w9","SWE_w10", "SWE_STMF")


### Special run for extended post-intervention window ------------
# #Post-intervention window set to week 26 of 2023
# postwindow <- 202326
# prewindows <- c(201627)
# mypredictors <- c("full")
# dependents <- c("RTotal_weekly100K")

# Save the variables to an R object file
save(prewindows, mypredictors, dependents, swe_related_codes, file = "SCM_variables.RData")


## Load and merge data --------------------------------------------
load("Mortality_data.Rdata")
load("WB_data.Rdata")
WB_cols <- setdiff(names(WB_data), c("CountryCode", "Year"))


# Function to fill and interpolate missing values within each country
impute_missing_data <- function(data, column_name) {
  if (all(is.na(data[[column_name]]))) {
    # If all values are missing, return the data as is
    return(data)
  } else {
    data %>%
      arrange(Year) %>%
      mutate(!!sym(column_name) := na.locf(!!sym(column_name), na.rm = FALSE)) %>% # Fill forward
      mutate(!!sym(column_name) := na.locf(!!sym(column_name), fromLast = TRUE)) %>% # Fill backward
      mutate(!!sym(column_name) := na.approx(!!sym(column_name), na.rm = FALSE)) # Interpolate
  }
}

WB_col <- WB_cols[1]

# Apply the function to each country/variable
for(wb in 1:length(WB_cols)){
  WB_col <- WB_cols[wb]
  WB_data <- WB_data %>%
    group_by(CountryCode) %>%
    group_modify(~ impute_missing_data(.x, WB_col)) %>%
    ungroup()
}

#Change Swedish country codes to SWE before merging data from WB on
df <- Mortality_data %>%
  mutate(
    CountryCode_org = CountryCode,  # Preserve original CountryCode
    CountryCode = substr(CountryCode,1,3)
  )

df <- left_join(df, WB_data, by=c("CountryCode", "Year"))

df <- df %>%
  mutate(CountryCode = CountryCode_org) %>%
  select(-CountryCode_org)

# Numeric CountryID
df <- df %>%
  mutate(CountryID = dense_rank(CountryCode))

df <- df %>%
  mutate(
    CountryID = as.numeric(factor(CountryCode)),
    YearWeek = Year * 100 + Week
      )


## Summarize data  ---------------------------------

### Summary of data availability --------------
# Filter the data to the specified YearWeek range
filtered_data <- df %>%
  filter(YearWeek >= 200727 & YearWeek <= 202126 & substr(CountryCode, 1, 4) != "SWE_")

# Identify countries that have data for both 200727 and 202126
valid_countries <- filtered_data %>%
  filter(YearWeek %in% c(202126)) %>%
  group_by(CountryCode) %>%
  summarize(count = n()) %>%
  filter(count == 1) %>%
  pull(CountryCode)

# Filter out countries that do not meet the criterion
filtered_data <- filtered_data %>%
  filter(CountryCode %in% valid_countries)


# Function to check if a country has complete data for a given year
has_complete_data <- function(data) {
  complete_data <- data %>%
    group_by(Year) %>%
    summarize(complete = all(!is.na(RTotal_weekly100K) & !is.na(R65p_weekly100K) &
                               !is.na(Hospital_beds) & !is.na(`Urban population`) & !is.na(GDPCAP_PPP) &
                               !is.na(ShareAged65p) & !is.na(Migrant_share)))
  complete_data
}

# Check data availability for each country
country_data_availability <- filtered_data %>%
  group_by(CountryCode) %>%
  do(has_complete_data(.))

# Summarize how many countries have complete data for each year
summary_data_availability <- country_data_availability %>%
  group_by(Year) %>%
  summarize(complete_countries = sum(complete))

# Print and save the summary table
print(summary_data_availability)
write.xlsx(summary_data_availability, file = paste0("Data availability.xlsx"))


## Define plot functions -----------------------------------------------------------

create_custom_plot <- function(df, lsw, min_ylim = NULL, max_ylim = NULL, yplace = NULL, ylabel = NULL) {
  # Find the position of 'lsw' within the sorted unique weeks
  all_weeks <- sort(unique(df$Sequence))
  lw_position <- match(lsw, all_weeks)
  
  # Generate sequences backwards and forwards from 'lsw', including 'lsw' itself
  myLabelCount <- ceiling(length(all_weeks) / 12)
  backward_indices <- seq(lw_position, 1, by = -myLabelCount)
  forward_indices <- seq(lw_position, length(all_weeks), by = myLabelCount)
  
  # Combine the sequences and remove duplicates
  combined_indices <- unique(c(backward_indices, forward_indices))
  combined_indices <- sort(combined_indices)  # Ensure it's sorted
  
  # Select the custom breaks and labels using the combined indices
  custom_breaks <- all_weeks[combined_indices]
  custom_labels <- unique(df$Year_w_Week[match(custom_breaks, df$Sequence)])
  
  # Set Y-scale
  if (is.null(max_ylim)) {
    max_outcome <- max(df$Outcome)
    max_ylim <- ceiling(max_outcome)
  }
  
  if (is.null(min_ylim)) {
    min_outcome <- min(df$Outcome)
    min_ylim <- floor(min_outcome)
  }
  
  if (is.null(yplace)) {
    yplace <- 0
  }
  
  if (is.null(ylabel)) {
    ylabel <- "Mortality/100,000"
  }
  
  # Plotting
  MyPlot <- 
    ggplot(df, aes(x = Sequence, y = Outcome, linetype = Type)) +
    geom_line() +
    theme_minimal() +
    theme(plot.background = element_rect(fill = "white"),  # Set plot background to white
          panel.background = element_rect(fill = "white"),  # Set panel background to white
          legend.position = "bottom",
          axis.text.x = element_text(angle = 90, hjust = 1)) + # Rotate x-axis labels
    scale_x_continuous(name = NULL, breaks = custom_breaks, labels = custom_labels) +
    scale_color_manual(values = rep("black", length(unique(df$Type)))) +  # Ensure one color per Type
    scale_linetype_manual(values = c("solid", "dotted")) +
    labs(title = NULL,
         y = ylabel,
         linetype  = NULL) +  # Removes default legend title
    geom_vline(xintercept = lsw, linetype = "dashed", color = "black", size = 1, show.legend = FALSE) +
    annotate("text", x = lsw - myLabelCount / 4, y = yplace, label = "Lockdown week", size = 4, color = "black", angle = 90, hjust = 0) +
    ylim(min_ylim, max_ylim) +
    geom_hline(yintercept = 0, linetype = "solid", color = "black", size = 0.8)  # Add horizontal line at y = 0
  
  return(MyPlot)
}

create_placebo_plot <- function(df, lsw, main, min_ylim = NULL, max_ylim = NULL, yplace = NULL) {
  # Find the position of 'lsw' within the sorted unique weeks
  all_weeks <- sort(unique(df$Sequence))
  lw_position <- match(lsw, all_weeks)
  
  # Generate sequences backwards and forwards from 'lsw', including 'lsw' itself
  myLabelCount <- ceiling(length(all_weeks) / 12)
  backward_indices <- seq(lw_position, 1, by = -myLabelCount)
  forward_indices <- seq(lw_position, length(all_weeks), by = myLabelCount)
  
  # Combine the sequences and remove duplicates
  combined_indices <- unique(c(backward_indices, forward_indices))
  combined_indices <- sort(combined_indices)  # Ensure it's sorted
  
  # Select the custom breaks and labels using the combined indices
  custom_breaks <- all_weeks[combined_indices]
  custom_labels <- unique(df$Year_w_Week[match(custom_breaks, df$Sequence)])
  
  # Set Y-scale
  if (is.null(max_ylim)) {
    max_outcome <- max(df$Outcome)
    max_ylim <- ceiling(max_outcome)
  }
  
  if (is.null(min_ylim)) {
    min_outcome <- min(df$Outcome)
    min_ylim <- floor(min_outcome)
  }
  
  if (is.null(yplace)) {
    yplace <- 0
  }
  
  # Add a group column to differentiate the first line from the rest
  df$LineGroup <- ifelse(grepl("^SWE", df$Type), main, "Placebos")
  
  # Define line sizes
  line_sizes <- c(setNames(0.6, main), "Placebos" = 0.3)
  
  # Plotting
  MyPlot <- 
    ggplot(df, aes(x = Sequence, y = Outcome, color = LineGroup, group = Type)) +
    geom_line(aes(linetype = LineGroup, size = LineGroup)) +
    theme_minimal() +
    theme(plot.background = element_rect(fill = "white"),  # Set plot background to white
          panel.background = element_rect(fill = "white"),  # Set panel background to white
          legend.position = "bottom",
          axis.text.x = element_text(angle = 90, hjust = 1)) + # Rotate x-axis labels
    scale_x_continuous(name = NULL, breaks = custom_breaks, labels = custom_labels) +
    scale_color_manual(values = c(setNames("black", main), "Placebos" = "grey")) +  # Set colors
    scale_linetype_manual(values = c(setNames("solid", main), "Placebos" = "solid")) +  # Ensure all lines are solid
    scale_size_manual(values = line_sizes) +  # Set line sizes
    labs(title = NULL,
         y = "Cumulative Excess Mortality/100,000",
         linetype = NULL,
         color = NULL,
         size = NULL) +  # Removes default legend title
    geom_vline(xintercept = lsw, linetype = "dashed", color = "black", size = 1, show.legend = FALSE) +
    annotate("text", x = lsw - myLabelCount / 4, y = yplace, label = "Lockdown week", size = 4, color = "black", angle = 90, hjust = 0) +
    ylim(min_ylim, max_ylim) +
    geom_hline(yintercept = 0, linetype = "solid", color = "black", size = 0.8)  # Add horizontal line at y = 0
  
  MyPlot
  return(MyPlot)
}

# ..............................................................----
# Define scenarios and prepare data ---------------------------------

# Set lockdown week
lw <- 202011

# LOOPS -----------------------
##Loop through all SCM scenarios................ --------------------------------------------------------------------

### d-loop ... (RTotal, R0_64, or R65p) ----
for (d in 1:length(dependents)){  #start of d-loop
  #Choose outcome variable - age group - (and predictor) for SCM 
  dependent <- dependents[d]
  dependent_folder_name <- sub("_weekly100K", "", dependent)

### p-loop ... (full or single) ----  
for (p in 1:length(mypredictors)){
  #define predictor sets
  predictor_list <- list()
  predictor_list[["full"]] <- c(dependent, WB_cols)  #full
  predictor_list[["single"]] <- dependent  #single
  
  mypredictor <- mypredictors[p]
  
  predictors <- predictor_list[[mypredictor]]
  
### pw-loop (pre-intervention window)... (200726, 201127, or 201627) ----
for (pw in 1:length(prewindows)){  #start of pw-loop

#### Create folders ----
  #Choose when pre-intervention window starts
  postyear <- floor(postwindow/100)
  prewindow <- prewindows[pw]
  
  # Define the folder names
  agegroup_folder <- paste0("Results ", postyear," - Agegroup '", dependent_folder_name, "'")
  predictor_folder <- paste0(agegroup_folder, "/Predictor '", mypredictor, "'")
  results_folder <- paste0(predictor_folder, "/Prewindow ", prewindow)
  
  # Check if the folder exists; if not, create it
  if (!dir.exists(agegroup_folder)) {
    dir.create(agegroup_folder)
  }

  if (!dir.exists(predictor_folder)) {
    dir.create(predictor_folder)
  }
  
  if (!dir.exists(results_folder)) {
    dir.create(results_folder)
  }

  

#### Filter data ----
  # Filter the dataset to keep relevant countries with a full mortality dataset
  filtered_df <- df %>%
    # Filter based on the prewindow and postwindow
    filter(YearWeek >= prewindow & YearWeek <= postwindow) %>%
    # Group by CountryCode to calculate min and max YearWeek per country
    group_by(CountryCode) %>%
    # Filter out countries that do not cover the full YearWeek range
    filter(min(YearWeek) == prewindow & max(YearWeek) == postwindow) %>%
    # Ungroup to return to the original dataframe structure
    ungroup()
  
  # Filter the dataset to keep relevant countries with a full WB dataset pre-intervention
  # Identify countries with NAs before the specified week number
  countries_with_na <- filtered_df %>%
    filter(YearWeek < lw & substr(CountryCode, 1, 3) != "SWE") %>%
    group_by(CountryCode) %>%
    summarise(has_na = any(across(everything(), ~ any(is.na(.))))) %>%
    filter(has_na) %>%
    pull(CountryCode)
  
  # Filter out those countries from the original data frame which do not have a full data set
  filtered_df <- filtered_df %>%
    filter(!CountryCode %in% countries_with_na)
  print(paste0(countries_with_na, " was dropped from the donor pool because of missing WB data."))
  
  # Create Sequence
  filtered_df <- filtered_df %>%
    group_by(CountryCode) %>%
    arrange(CountryCode, YearWeek) %>% # Ensure the data is sorted by YearWeek within each country
    mutate(Sequence = row_number()) %>%
    ungroup()
  
  # Create table for use later
  week_to_yearweek <- unique(filtered_df[c("Sequence", "YearWeek")])
  week_to_yearweek <- week_to_yearweek %>%
    mutate(Year_Week = paste(substr(YearWeek, 1, 4), substr(YearWeek, 5, 6), sep = "-"),
           Year_w_Week = paste(substr(YearWeek, 1, 4), substr(YearWeek, 5, 6), sep = "w"))
  
#### Set variables for the SCM --------------------------------------

  #Lockdown sequence week
  lsw <- filtered_df %>% filter(YearWeek == lw) %>% pull(Sequence) %>% max()  # Using 'pull' to directly extract the column values

if(!onlyPlots){   
  #number of weeks
  FinalWeek <- max(filtered_df$Sequence)
  
#### Define donor pool and data -----------
  Donordata <- filtered_df %>%
    # Filter out unnecessary Sweden-versions first
    filter(!(CountryCode %in% swe_related_codes)) %>%
    # Filter based on the prewindow and postwindow
    filter(YearWeek >= prewindow & YearWeek <= postwindow) %>%
    # Group by CountryCode to calculate min and max YearWeek per country
    group_by(CountryCode) %>%
    # Filter out countries that do not cover the full YearWeek range
    filter(min(YearWeek) == prewindow & max(YearWeek) == postwindow) %>%
    # Ungroup to return to the original dataframe structure
    ungroup()
  
  Donordata <- as.data.frame(Donordata)
  
  controlsidentifier <- Donordata %>%
    pull(CountryID) %>%
    unique()
  
  # Setup for the placebo test
  placebo_plot_data_cum_list <- list()
  placebo_plot_data_list <- list()

#### Create placebo test data by looping through all controls ------------------------
  # Loop over each control unit to perform placebo tests
  for (j in 1:length(controlsidentifier)) {
    treated <- controlsidentifier[j]
    treated_cc <- Donordata %>% 
      filter(CountryID == treated) %>%
      pull(CountryCode) %>%
      unique()

    # Prepare data for the current control unit as if it were the treated unit
    dataprep_placebo <- dataprep(
      foo = Donordata,
      predictors = predictors,
      predictors.op = "mean",
      dependent = dependent,
      unit.variable = "CountryID",
      time.variable = "Sequence",
      unit.names.variable = "CountryCode",
      treatment.identifier = treated,
      controls.identifier = setdiff(controlsidentifier, treated),
      time.predictors.prior = c(1:lsw),
      time.optimize.ssr = c(1:lsw),
      time.plot = c(1:FinalWeek)
    )
    
    # Run synth for the placebo
    synth_placebo <- synth(data.prep.obj = dataprep_placebo, optimxmethod = "All")
    
    # Calculate the synthetic control for the placebo
    synth_placebo_control <- dataprep_placebo$Y0plot %*% synth_placebo$solution.w
    
#### Create cumulative graphs data  --------------------------------------------------------------
    
    # Graphs data
    plot_data <- data.frame(
      Sequence = 1:FinalWeek,
      Actual = Donordata[[dependent]][Donordata$CountryID == treated],  # Use dynamic column name
      Synthetic = as.numeric(synth_placebo_control)  # Data for synthetic control
    )
    
    # Create data for cumulative effect
    plot_data_cum <- plot_data %>% 
      mutate(
        `Weekly Effect` = ifelse(Sequence <= lsw, 0, Actual - Synthetic),
        `Cumulative Effect` = cumsum(`Weekly Effect`)
      ) %>%
      select(Sequence, `Cumulative Effect`) %>%
      rename_with(~ treated_cc, `Cumulative Effect`)
    
    # Melting the data frame for ggplot2
    plot_data_long_cum <- reshape2::melt(plot_data_cum, id.vars = "Sequence", variable.name = "Type", value.name = "Outcome")
    
    # Store results
    placebo_plot_data_cum_list[[treated_cc]] <- plot_data_long_cum
    placebo_plot_data_list[[treated_cc]] <- plot_data
  }
#### Save placebo_data ----
  save(placebo_plot_data_cum_list, file = paste0(results_folder, "/placebo_plot_data_cum_list.RData"))
  save(placebo_plot_data_list, file = paste0(results_folder, "/placebo_plot_data_list.RData"))
} #end if(onlyPlots)

#### Create containers for data ------------
  dfWeights <- data.frame()
  plot_data_long_list <- list()
  plot_data_list <- list()
  plot_data_cum_list <- list()

  
## i-loop ... (SWE, SWE_w7 etc.) ----------

if(onlyPlots){
  # Delete all plots in result_folder
  unlink(paste0(results_folder,"/*.png"))
}


  
for (i in 1:length(swe_related_codes)) {
  #Skip irrelevant combinations
  if (!((pw %in% 1:3 & i == 1) | (pw == 1 & i %in% 4:7))) {
    next  # Skip the combination if it doesn't match the criteria
  }
  
  
  # Select the SWE-related country code you want to analyze
  MySce <- swe_related_codes[i]
  
if(!onlyPlots){
## Create SCMdata ---------------------------------------------------------
  
  ## Filter the dataset to keep the relevant Sweden
  SWEdata <- filtered_df %>% filter(CountryCode == MySce)
  
  treatmentidentifier <- SWEdata %>%
    pull(CountryID) %>%
    unique()
  
  SCMdata <- bind_rows(Donordata, SWEdata)
  
  SCMdata <- as.data.frame(SCMdata)
  
  #### QA of SCMdata ---------------------------------
  # Check that the data are balanced
  summary <- SCMdata %>%
    group_by(CountryCode) %>%
    summarize(
      Num_Obs = n(),
      .groups = "drop"
    )
  if(min(summary$Num_Obs) != max(summary$Num_Obs)){stop("The data are unbalanced")}

### Create special predictors to place greater emphasis on the most recent development ---------------------------------------------------------
  # Identify the sequence numbers corresponding to week 27 of 2018 and week 27 of 2019
  start_emphasis_sequence_2018 <- SCMdata$Sequence[SCMdata$Year == 2018 & SCMdata$Week == 27][1]
  start_emphasis_sequence_2019 <- SCMdata$Sequence[SCMdata$Year == 2019 & SCMdata$Week == 27][1]

### Create special predictors to place emphasis on the development before lockdowns can possible be effective ---------------------------------------------------------
  post_intervention_week1 <- lsw + 1
  post_intervention_week2 <- lsw + 2


## Run SCM ---------------------------------------------------------------------

  dataprep <-
    dataprep(
      foo = SCMdata,
      predictors = predictors,
      predictors.op = "mean",
      dependent     = dependent,
      unit.variable = "CountryID",
      time.variable = "Sequence",
      unit.names.variable   = "CountryCode",
      treatment.identifier  = treatmentidentifier,
      controls.identifier   = controlsidentifier,
      time.predictors.prior = c(1:lsw),
      time.optimize.ssr     = c(1:lsw),
      # special.predictors are used as a robustness check
      # special.predictors    = list(
      #   list(dependent, start_emphasis_sequence_2018:lsw, c("mean")), # Emphasize the period from week 27 of 2018 onward
      #   list(dependent, start_emphasis_sequence_2019:lsw, c("mean")), # Emphasize the period from week 27 of 2019 onward
      #   list(dependent, post_intervention_week1, c("mean")),          # Include the first week after the intervention
      #   list(dependent, post_intervention_week2, c("mean"))          # Include the second week after the intervention
      # ),
      time.plot            = c(1:FinalWeek)
    )
  
  # run synth
  synth.out <- synth(data.prep.obj = dataprep, optimxmethod="All")
  
  synth.SWE<-(dataprep$Y0plot%*%synth.out$solution.w)


#### Create plot data --------------------------------------------------------------
  ##### Weekly effect -------------------
  
  # Graphs data
  plot_data_org <- data.frame(
    Sequence = 1:FinalWeek,
    Actual = SCMdata[[dependent]][SCMdata$CountryID == treatmentidentifier],  # Use dynamic column name
    Synthetic = as.numeric(synth.SWE)  # Data for synthetic control
  )
  
  # #Normalize synthetic to average of 201927 to 202011
  # norm_factor <- plot_data_org %>%
  #   filter(Sequence >= lsw-26-11 & Sequence <=lsw) %>%
  #   mutate(diff = Synthetic -Actual) %>%
  #   select(diff) %>%
  #   unlist() %>%
  #   mean()
  # 
  # plot_data <- plot_data_org %>%
  #   mutate(Synthetic = Synthetic - norm_factor)
  
  plot_data <- plot_data_org
  
  # Store plot_data
  plot_data_list[[MySce]] <- plot_data
  
  # Melting the data frame for ggplot2
  plot_data_long <- reshape2::melt(plot_data, id.vars = "Sequence", variable.name = "Type", value.name = "Outcome")
  
  # Add more options to the x-axis
  plot_data_long <- merge(plot_data_long, week_to_yearweek, by = "Sequence", all.x = TRUE)

  # Store plot_data_long
  plot_data_long_list[[MySce]] <- plot_data_long
  
  ##### Cumulative effect -------------------------
  
  # Create data for cumulative effect
  plot_data_cum <- plot_data %>%
    # Calculate the Weekly difference
    mutate(Weekly = Actual - Synthetic) %>%
    # Calculate the non-standardized cumulative sum
    mutate(Cumulative_nonstandardized = cumsum(Weekly)) %>%
    # Extract the value of Cumulative_nonstandardized at lsw
    mutate(LSW_Cumulative = ifelse(Sequence == lsw, Cumulative_nonstandardized, NA)) %>%
    # Carry forward and backward the LSW_Cumulative value
    mutate(LSW_Cumulative = zoo::na.locf(zoo::na.locf(LSW_Cumulative, na.rm = FALSE), fromLast = TRUE)) %>%
    # Normalize the Cumulative_nonstandardized by subtracting the value at lsw
    mutate(Cumulative = Cumulative_nonstandardized - LSW_Cumulative) %>%
    # Select relevant columns
    select(Sequence, Cumulative, Weekly)
  
  # Melting the data frame for ggplot2
  plot_data_long_cum <- reshape2::melt(plot_data_cum, id.vars = "Sequence", variable.name = "Type", value.name = "Outcome")
  
  # Add more options to the x-axis
  plot_data_long_cum <- merge(plot_data_long_cum, week_to_yearweek, by = "Sequence", all.x = TRUE)
  
  # Store plot_data_long_cum
  plot_data_cum_list[[MySce]] <- plot_data_long_cum
  
  
  #### Generate tables ---------------------------------------------------------
  
  # Creates table with weights
  synth.tables <- synth.tab(
    dataprep.res = dataprep,
    synth.res = synth.out
  )
  
  # Rename 'w.weights' to the value stored in MySce, drop 'unit.numbers' and left-join
  if (nrow(dfWeights) == 0) {
    dfWeights <- synth.tables$tab.w %>%
      select(Country := unit.names, !!sym(MySce) := w.weights, -unit.numbers)
  } else {
    # Remove the existing column if it exists
    if (MySce %in% colnames(dfWeights)) {
      dfWeights <- dfWeights %>%
        select(-all_of(MySce))
    }
    
    # Perform the left join to add the new data
    dfWeights <- dfWeights %>%
      left_join(
        synth.tables$tab.w %>%
          select(Country := unit.names, !!sym(MySce) := w.weights),
        by = "Country"
      )
  }

} #end if(!onlyPlots)
## ##Loop through all plot creations................ ------------------------------------------

### Prepare data  -----
if(onlyPlots){
  #Load plot_data_long
  load(paste0(results_folder, "/plot_data_long_list.RData"))
  plot_data_long <- plot_data_long_list[[MySce]]
  
  #Load plot_data_long_cum
  load(paste0(results_folder, "/plot_data_cum_list.RData"))
  plot_data_long_cum <- plot_data_cum_list[[MySce]] 
  
  #Load placebo_plot_data_cum_list
  load(paste0(results_folder, "/placebo_plot_data_cum_list.RData"))
}
### Fix plot data and set limits ----
  #Fix label for graphs
  mylabel <- sub("SWE_?", "Sweden", MySce)  #Replace SWE and SWE_ with Sweden
  mylabel <- sub("STMF\\b", " (STMF)", mylabel)
  mylabel <- sub("w7\\b", " (w7)", mylabel)
  mylabel <- sub("w8\\b", " (w8)", mylabel)
  mylabel <- sub("w9\\b", " (w9)", mylabel)
  mylabel <- sub("w10\\b", " (w10)", mylabel)
  mylabel <- sub("w78\\b", " (w7+w8)", mylabel)
  mylabel <- sub("w910\\b", " (w9+w10)", mylabel)
  MyNum <- i
  
  # Set file name info
  plot_agegroup <- dependent_folder_name
  plot_predictor <- mypredictor
  plot_prewindow <- substr(prewindow, 1, 4)
  plot_info <- paste(plot_agegroup, plot_prewindow ,plot_predictor, sep="_")
  
  
  plot_data_long <- plot_data_long %>%
    mutate(Type = ifelse(Type == "Synthetic", paste0("Synthetic ", mylabel), paste0("Actual ", mylabel)))
  
  ymin <- floor(min(plot_data_long$Outcome))
  ymax <- ceiling(max(plot_data_long$Outcome))
  
  yp <- plot_data_long %>% 
    filter(Sequence <= lsw & Sequence >= lsw-52) %>%
    select(Outcome) %>%
    max()
  
  yp <- yp+2
  
### Plot with full pre- and post-intervention window ---------
  MyPlot <- create_custom_plot(plot_data_long, lsw, min_ylim = ymin, max_ylim = ymax, yplace = yp)
  MyPlot
  ggsave(paste0(results_folder, "/Plot_", MySce, "_", plot_info, ".png"), plot = MyPlot, width = 5, height = 5, dpi = 300, bg = "white")
  ggsave(paste0(results_folder, "/Wide_", MySce, "_", plot_info, ".png"), plot = MyPlot, width = 10, height = 5, dpi = 300, bg = "white")
  
### Plot focusing on post-intervention window (zoom)------------
  plot_data_long_zoom <- plot_data_long[plot_data_long$YearWeek >= 202001,]
  MyPlotZoom <- create_custom_plot(plot_data_long_zoom, lsw, min_ylim = ymin, max_ylim = ymax, yplace = yp)
  MyPlotZoom
  ggsave(paste0(results_folder, "/Zoom_", MySce, "_", plot_info, ".png"), plot = MyPlotZoom, width = 5, height = 5, dpi = 300, bg = "white")
  
### Cum plot focusing on effect & cumulative effect post-intervention ------------
  yp <- floor(min(plot_data_long_cum$Outcome)) 
  
  MyPlotCum <- create_custom_plot(plot_data_long_cum, lsw, yplace = yp, ylabel = "Excess Mortality/100,000")
  MyPlotCum
  ggsave(paste0(results_folder, "/Cum_", MySce, "_", plot_info, ".png"), plot = MyPlotCum, width = 5, height = 5, dpi = 300, bg = "white")
  
  
### Cum plot focusing on post-intervention window (zoom) ------------
  plot_data_long_cum_zoom <- plot_data_long_cum %>%
    filter(YearWeek >= 202001) %>%
    mutate(Outcome = ifelse(YearWeek <= lw, 0, Outcome))
  
  yp <- floor(min(plot_data_long_cum_zoom$Outcome)) 
  
  MyPlotCumZoom <- create_custom_plot(plot_data_long_cum_zoom, lsw, yplace = yp, ylabel = "Excess Mortality/100,000")
  MyPlotCumZoom
  ggsave(paste0(results_folder, "/CumZoom_", MySce, "_", plot_info, ".png"), plot = MyPlotCumZoom, width = 5, height = 5, dpi = 300, bg = "white")

  
  
### Placebo test plot ------------------------------------------------------
  
  # Bind the filtered results
  plot_data_long_placebo <- bind_rows(placebo_plot_data_cum_list)
  
  # Add WeekNum etc. to the 
  plot_data_long_placebo <- merge(plot_data_long_placebo, week_to_yearweek, by = "Sequence", all.x = TRUE)
  
  #Add the effect from our main scenario
  plot_data_long_main <- plot_data_long_cum %>%
    filter(Type == "Cumulative") %>%
    mutate(Type = MySce)
  
  # Update plot_data_long_placebo with main first
  plot_data_long_placebotest <- bind_rows(plot_data_long_main, plot_data_long_placebo)
  
  # Filter to period with an effect (YearWeek >= )
  plot_data_long_placebotest <- plot_data_long_placebotest %>% filter(YearWeek >= 202001)
  
  MyPlotPlacebo <- create_placebo_plot(plot_data_long_placebotest, lsw, MySce)
  MyPlotPlacebo
  ggsave(paste0(results_folder, "/Placebo_",MySce, "_", plot_info, ".png"), plot = MyPlotPlacebo, width = 5, height = 5, dpi = 300, bg = "white")    

##End loops ----
} ###End i-loop ----

if(!onlyPlots){
### Save weights and plot data ----------------------------------------------
  write.xlsx(dfWeights, file = paste0(results_folder, "/Weights.xlsx"))
  save(plot_data_list, file = paste0(results_folder, "/plot_data_list.RData"))
  save(plot_data_long_list, file = paste0(results_folder, "/plot_data_long_list.RData"))
  save(plot_data_cum_list, file = paste0(results_folder, "/plot_data_cum_list.RData"))
}
  save(week_to_yearweek, file = paste0(results_folder, "/week_to_yearweek.RData"))
} ###End of pw-loop ----
} ###End of p-loop ----
} ###End of d-loop ----


# Extra calculations showing problems with SCM------------------------------------------------------

#Load plot_data_long
load("Results 2021 - Agegroup 'RTotal'/Predictor 'full'/Prewindow 201627/plot_data_long_list.RData")
plot_data_long_w7 <- plot_data_long_list[["SWE_w7"]] %>% filter(YearWeek >= 202001) 
plot_data_long_w8 <- plot_data_long_list[["SWE_w8"]] %>% filter(YearWeek >= 202001)
plot_data_long_w9 <- plot_data_long_list[["SWE_w9"]] %>% filter(YearWeek >= 202001)
plot_data_long_w10 <- plot_data_long_list[["SWE_w10"]] %>% filter(YearWeek >= 202001)

df <- bind_rows(plot_data_long_w7 %>% filter(Type == "Synthetic") %>% mutate(Type = "w7"),
                                       plot_data_long_w8 %>% filter(Type == "Synthetic") %>% mutate(Type = "w8"),
                                       plot_data_long_w9 %>% filter(Type == "Synthetic") %>% mutate(Type = "w9"),
                                       plot_data_long_w10 %>% filter(Type == "Synthetic") %>% mutate(Type = "w10"))

lsw <- 193
min_ylim <- NULL
max_ylim <- NULL
yplace <- NULL
ylabel <- NULL


## Plot --------------------------------------------------------------------


# create_four_lines_plot <- function(df, lsw, min_ylim = NULL, max_ylim = NULL, yplace = NULL, ylabel = NULL) {
# Find the position of 'lsw' within the sorted unique weeks
all_weeks <- sort(unique(df$Sequence))
lw_position <- match(lsw, all_weeks)

# Generate sequences backwards and forwards from 'lsw', including 'lsw' itself
myLabelCount <- ceiling(length(all_weeks) / 12)
backward_indices <- seq(lw_position, 1, by = -myLabelCount)
forward_indices <- seq(lw_position, length(all_weeks), by = myLabelCount)

# Combine the sequences and remove duplicates
combined_indices <- unique(c(backward_indices, forward_indices))
combined_indices <- sort(combined_indices)  # Ensure it's sorted

# Select the custom breaks and labels using the combined indices
custom_breaks <- all_weeks[combined_indices]
custom_labels <- unique(df$Year_w_Week[match(custom_breaks, df$Sequence)])

# Set Y-scale
if (is.null(max_ylim)) {
  max_outcome <- max(df$Outcome)
  max_ylim <- ceiling(max_outcome)
}

if (is.null(min_ylim)) {
  min_outcome <- min(df$Outcome)
  min_ylim <- floor(min_outcome)
}

if (is.null(yplace)) {
  yplace <- 0
}

if (is.null(ylabel)) {
  ylabel <- "Mortality/100,000"
}

my_colors <- rep("black", length(unique(df$Type)))

# Plotting
MyPlot <- 
  ggplot(df, aes(x = Sequence, y = Outcome, linetype = Type, color = Type)) +
  geom_line() +
  theme_minimal() +
  theme(plot.background = element_rect(fill = "white"),  # Set plot background to white
        panel.background = element_rect(fill = "white"),  # Set panel background to white
        legend.position = "bottom",
        axis.text.x = element_text(angle = 90, hjust = 1)) + # Rotate x-axis labels
  scale_x_continuous(name = NULL, breaks = custom_breaks, labels = custom_labels) +
  scale_color_manual(values = my_colors) +  # Ensure one color per Type
  scale_linetype_manual(values = c("solid", "dashed", "dotdash", "dotted")) +
  labs(title = NULL,
       y = ylabel,
       linetype  = NULL,
       color = NULL) +  # Removes default legend title
  geom_vline(xintercept = lsw, linetype = "dashed", color = "black", size = 1, show.legend = FALSE) +
  annotate("text", x = lsw - myLabelCount / 4, y = yplace, label = "Lockdown week", size = 4, color = "black", angle = 90, hjust = 0) +
  ylim(min_ylim, max_ylim) +
  geom_hline(yintercept = 0, linetype = "solid", color = "black", size = 0.8)  # Add horizontal line at y = 0

MyPlot

ggsave("Synths.png", plot = MyPlot, width = 5, height = 5, dpi = 300, bg = "white")
```
